# Modelling the cost-effectiveness of hepatitis A vaccination in South Africa

**DOI:** 10.1101/2022.10.03.22280656

**Authors:** J. Patterson, S. Cleary, H. van Zyl, T. Awine, S. Mayet, J. Norman, B. M. Kagina, R. Muloiwa, G. D. Hussey, S. P. Silal, Modelling the cost-effectiveness of hepatitis A vaccination in South Africa

## Abstract

The World Health Organization (WHO) recommends the consideration of introducing routine hepatitis A vaccination into national immunization schedules for children ≥ 1 years old in countries with intermediate HAV endemicity. Recent data suggest that South Africa is transitioning from high to intermediate HAV endemicity, thus it is important to consider the impact and cost of potential routine hepatitis A vaccination strategies in the country.

An age-structured compartmental model of hepatitis A transmission was calibrated with available data from South Africa, incorporating direct costs of hepatitis A treatment and vaccination. We used the calibrated model to evaluate the impact and costs of several childhood hepatitis A vaccination scenarios from 2023 to 2030. We assessed how each scenario impacted the burden of hepatitis A (symptomatic hepatitis A cases and mortality) as well as calculated the incremental cost per DALY averted as compared to the South African cost-effectiveness threshold. All costs and outcomes were discounted at 5%.

For the modelled scenarios, the median estimated cost of the different vaccination strategies ranged from $1.71 billion to $2.85 billion over the period of 2023 to 2030, with the cost increasing for each successive scenario and approximately 39-52% of costs being due to vaccination. Scenario 1, which represented the administration of one dose of the hepatitis A vaccine in children < 2 years old, requires approximately 5.3 million vaccine doses over 2023-2030 and is projected to avert a total of 136,042 symptomatic cases [IQR: 88,842-221,483] and 31,106 [IQR: 22,975-36,742] deaths due to hepatitis A over the period of 2023 to 2030. The model projects that Scenario 1 would avert 8,741 DALYs over the period of 2023 to 2030, however is not cost-effective against the South African cost-effectiveness threshold with an ICER per DALY averted of $21,006. While Scenario 3 and 4 included the administration of more vaccine doses and averted more symptomatic cases of hepatitis A, these scenarios were absolutely dominated owing to the population being infected before vaccination through the mass campaigns at older ages.

The model was highly sensitivity to varying access to liver transplant in South Africa. When increasing the access to liver transplant to 100% for baseline and Scenario 1, the ICER for Scenario 1 becomes cost-effective against the CET (ICER = $2,425). Given these findings, we recommend further research is conducted to understand the access to liver transplants in South Africa to better estimate the cost of liver transplant care for hepatitis A patients. The modelling presented in this paper has been used to develop a <u>user-friendly application</u> for vaccine policy makers to further interrogate the model outcomes and consider the costs and benefits of introducing routine hepatitis A vaccination in South Africa.

## 1.2 Background

Over the last two decades, South Africa has been assumed to have high hepatitis A virus (HAV) endemicity with seroprevalence ≥ 90% by 10 years old. Data suggests, however, that South Africa has transitioned from high to intermediate or low hepatitis A virus (HAV) endemicity with less children acquiring hepatitis A infection and developing natural immunity at a young age. With this shift and a rise in age of people susceptible to HAV infection in the population, the risk for serious outbreaks and significant burden of disease increases.

The World Health Organization (WHO) recommends the consideration of introducing routine hepatitis A vaccination into national immunization schedules for children ≥ 1 years old in countries with intermediate HAV endemicity. Previously published studies have found routine hepatitis A vaccination strategies to be cost-effective in countries with existing childhood immunization programs, however an analytical framework to assess the impact and cost of different routine hepatitis A vaccination strategies in South Africa has not yet been developed (1-10).

While the Expanded Program on Immunization in South Africa (EPI-SA) has been a leader in adopting new vaccines on the African continent, there are considerable economic obstacles facing the introduction of new vaccines into the EPI-SA. Implementation of new vaccines requires large upfront investment, and the success of new vaccination programs is often uncertain in LMICs. In countries with health budgets that have little room for expansion, it is important for economic evaluations to deliver strong evidence for opportunities of cost-effectiveness. We evaluated the cost, outcomes, cost-effectiveness of different potential routine hepatitis A vaccination strategies in South Africa. This model was developed with the aim to support the South African National Advisory Group on Immunization (NAGI) Hepatitis A Working Group’s consideration of introducing routine hepatitis A vaccination into the EPI-SA.

## 1.3 Methods

### 1.3.1 Transmission model

Ordinary differential equations were used to develop an age-structured model for hepatitis A transmission dynamics in South Africa. The model diagram is displayed in **Figure 5.1** and the differential equations are presented in **Supplementary Table S5.1**. In the model, the South African population is divided into 18 distinct hepatitis-A specific epidemiological compartments (**Table 5.1**), which are further stratified by 19 age groups (annual ages until 9 years old followed by 5-year age groups). The population is modelled over time through the birth rate, aging rate, and age-specific death rate.

**Figure 5.1:**
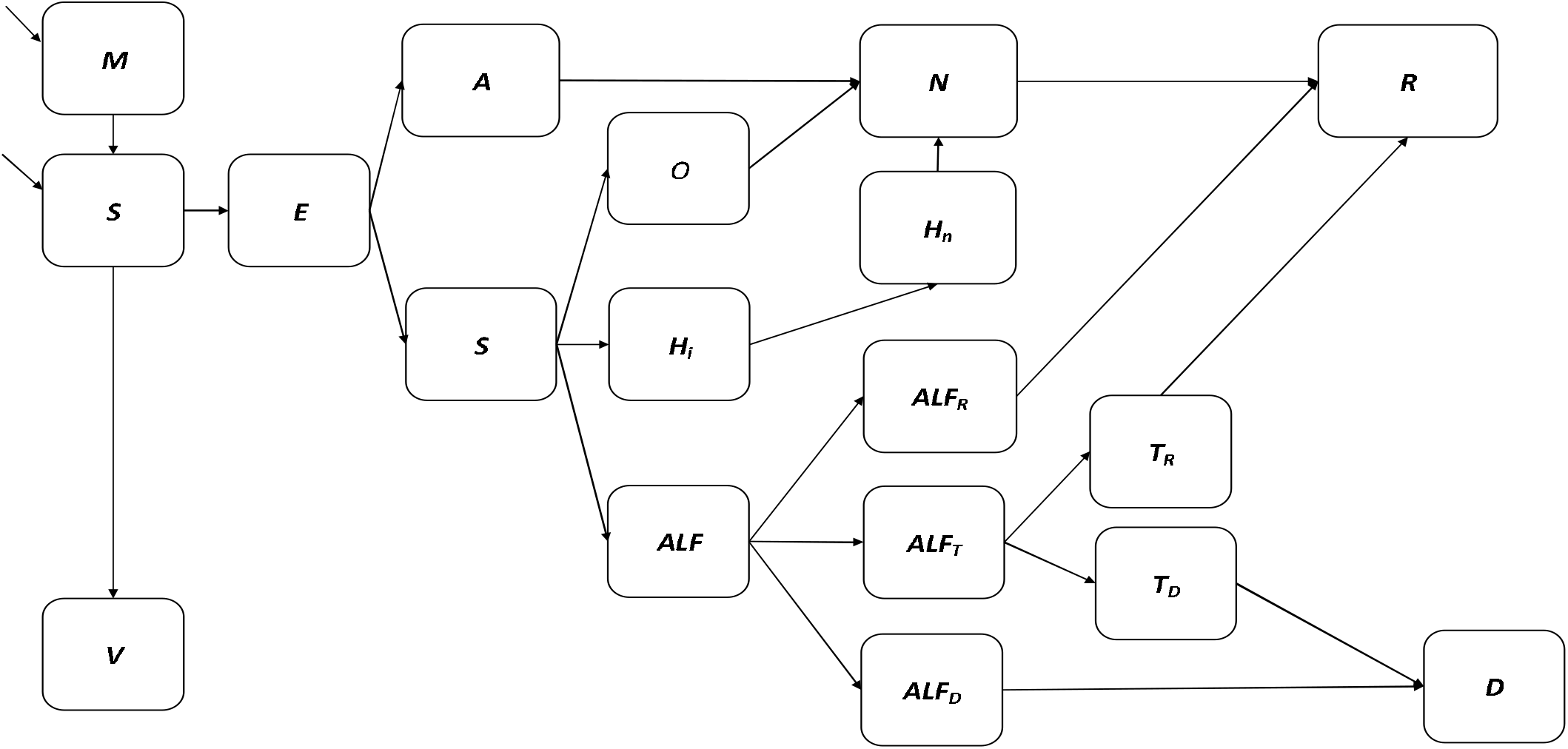
Flow diagram of hepatitis A transmission and vaccination model

**Figure 5.2:**
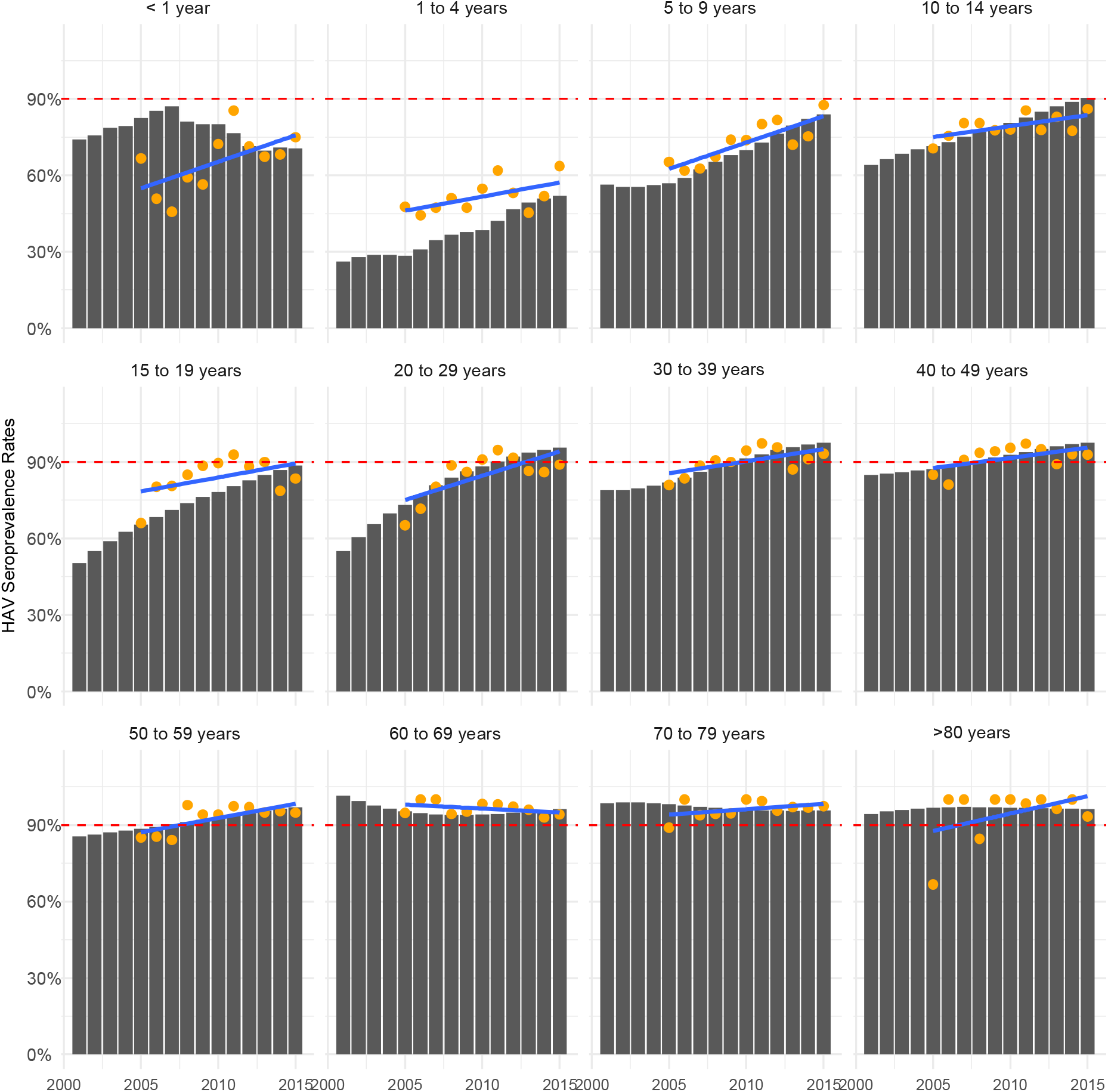
Model fitting to HAV seroprevalence (anti-HAV IgG) data by age group Legend Model output HAV seroprevalence data Trendline in HAV seroprevalence data

**Figure 5.3:**
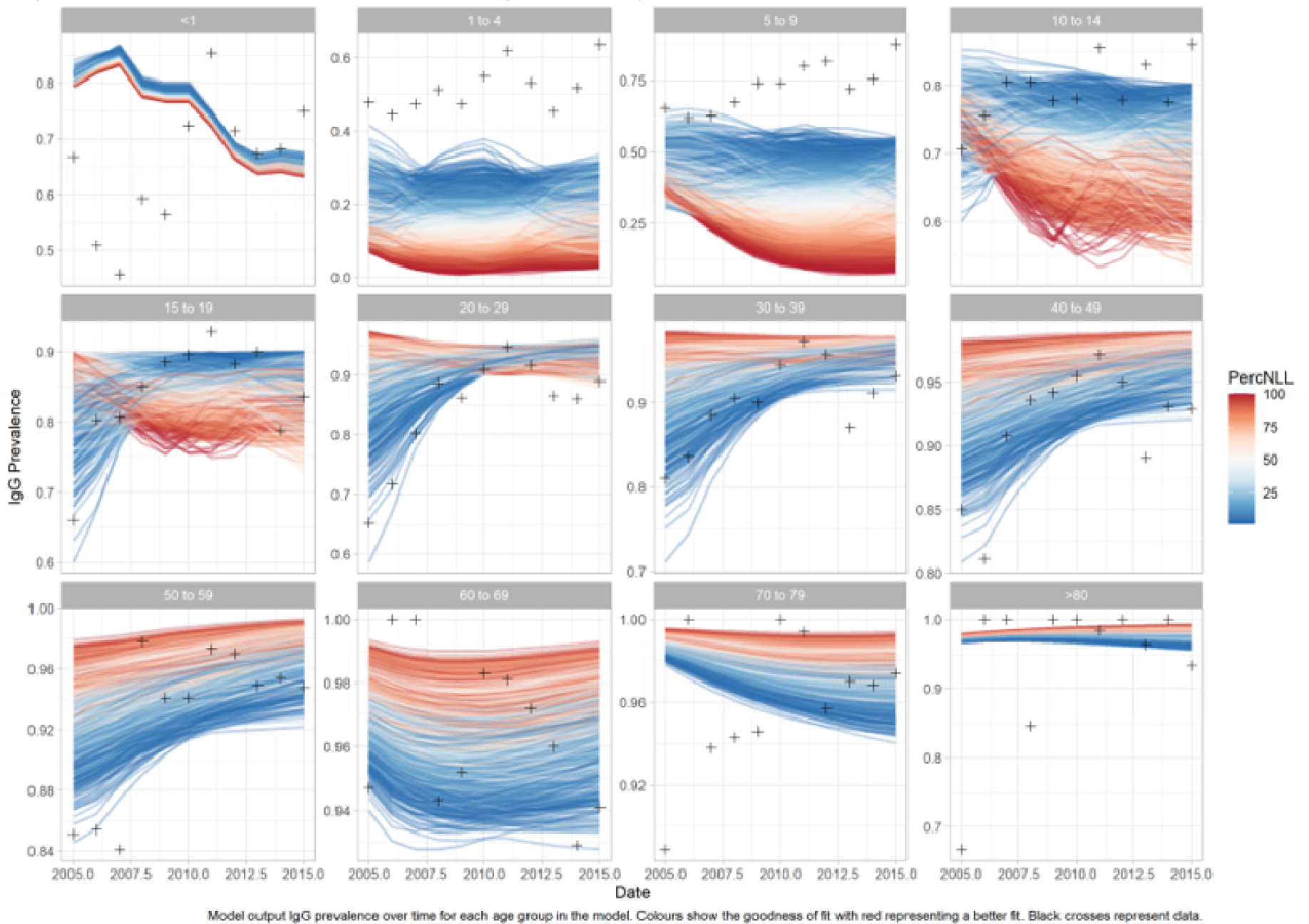
Model calibration negative log likelihood results

**Figure 5.4:**
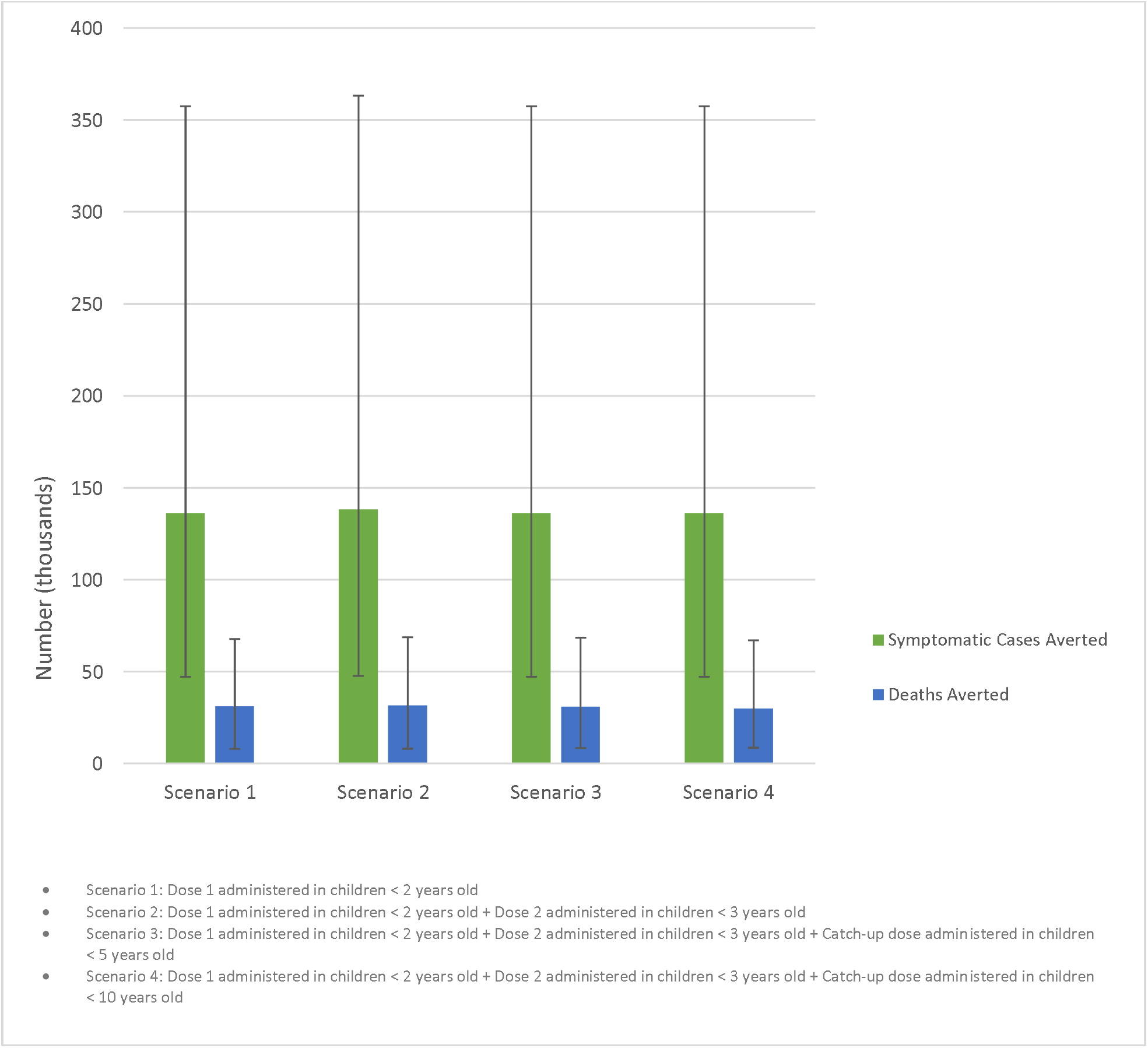
Impact of modelled vaccination scenarios on the burden of hepatitis A

**Figure 5.5:**
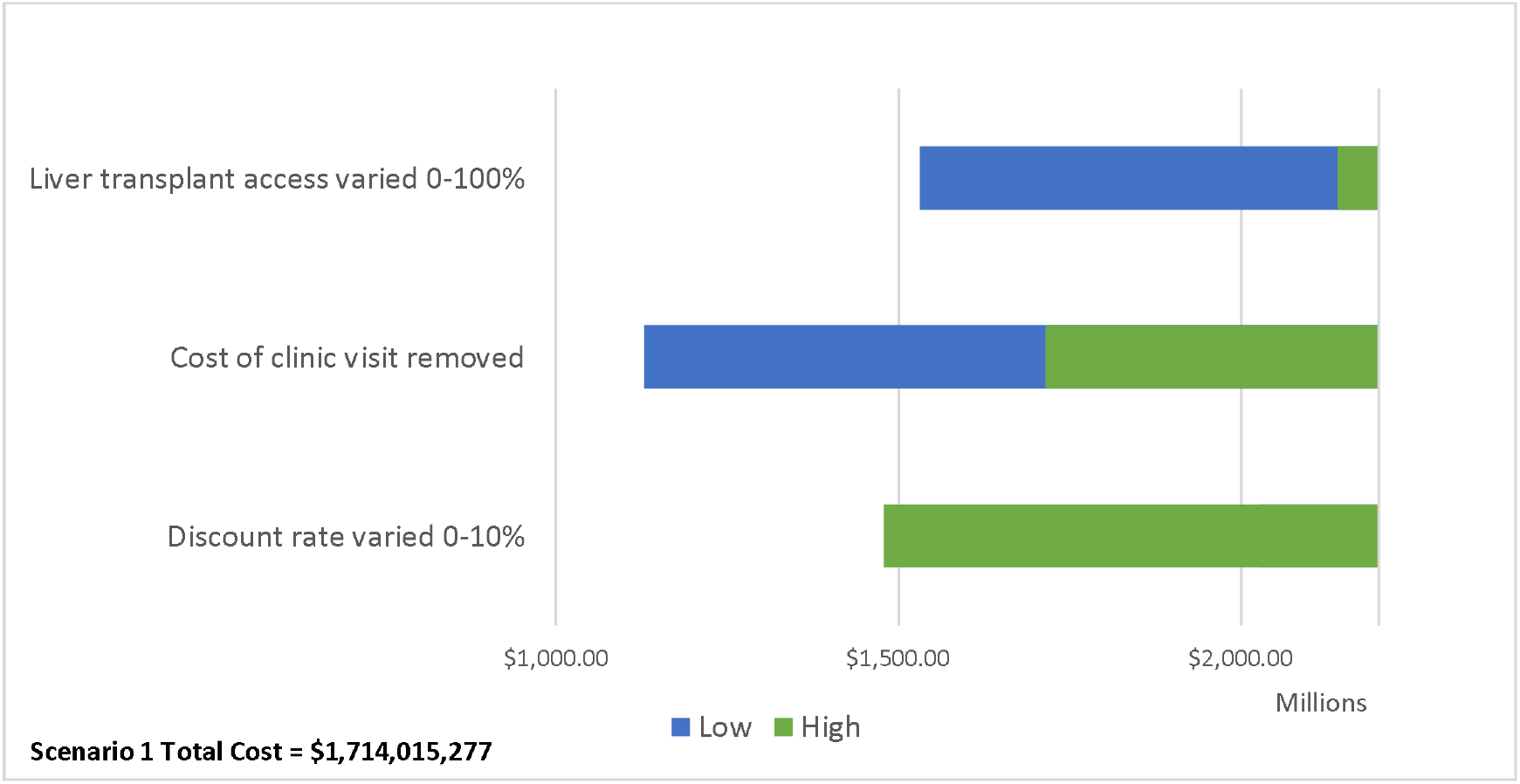
One-way sensitivity analysis impact on Scenario 1 total costs

**Table 5.1:** Model compartments and description

| Abbreviation | Compartment | Description |
| --- | --- | --- |
| M | Maternal antibodies | Presence of maternally acquired anti-HAV IgG antibodies |
| S | Susceptible | No presence of anti-HAV IgG antibodies |
| E | Exposed | Exposed to the hepatitis A virus with the risk of infection |
| A | Asymptomatic | Infected with the hepatitis A virus following the incubation period |
| S | Symptomatic | Infected with the hepatitis A virus following the incubation period |
| O | Outpatient case | Hepatitis A case requiring outpatient care at a tertiary level facility |
| H<sub>i</sub> | Hospitalized infectious case | Hepatitis A case requiring hospitalization at a tertiary level facility while shedding HAV |
| H<sub>n</sub> | Hospitalized non-infectious case | Hepatitis A case requiring hospitalization at a tertiary level facility after shedding HAV |
| N | Recovering case | Hepatitis A case with waning anti-HAV IgM antibodies |
| R | Recovered and immune | Previous hepatitis A case with anti-HAV IgG antibodies developed through infection |
| ALF | Viral-induced acute liver failure | Hepatitis A case resulting in acute liver failure defined as the development of encephalopathy and synthetic function impairment following acute liver injury in an individual without pre-existing liver disease |
| ALF<sub>R</sub> | Spontaneous recovery from acute liver failure | Viral-induced acute liver case that recovers without liver transplant |
| ALF<sub>D</sub> | Death due to acute liver failure | Viral-induced acute liver case that dies due to any cause |
| ALF<sub>T</sub> | Liver transplant case | Viral-induced acute liver transplant case that requires liver transplant for recovery |
| T<sub>R</sub> | Liver transplant recovery | Viral-induced acute liver transplant case that requires and receives liver transplant |
| T<sub>D</sub> | Liver transplant death | Liver transplant case that dies due to any cause |
| D | Hepatitis A death | Hepatitis A case that dies due to any cause |
| V | Vaccinated | Vaccinated with one or two doses of hepatitis A vaccine with sufficient development of anti-HAV IgG antibodies for protection against infection |

**Table 5.2:**
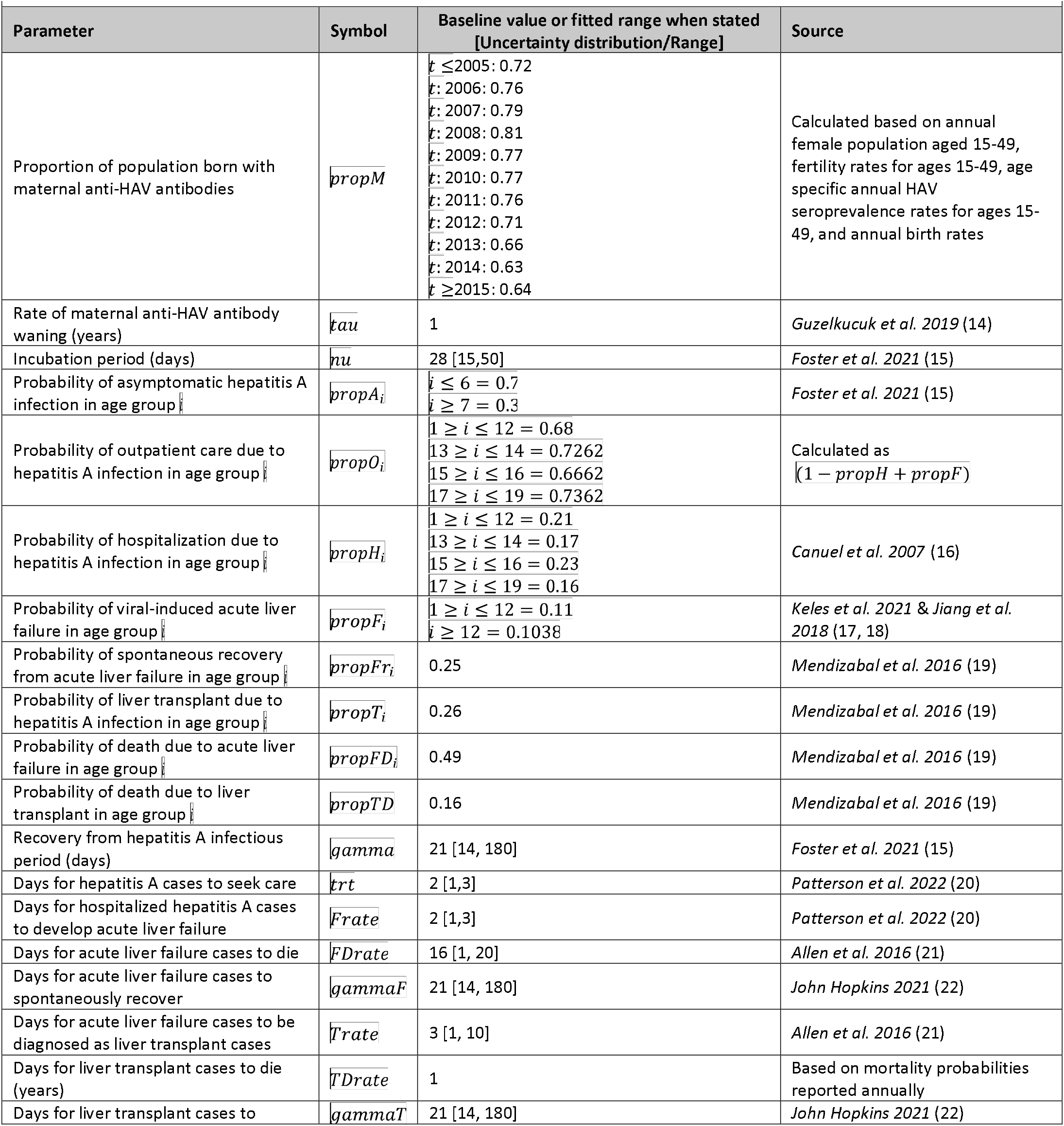
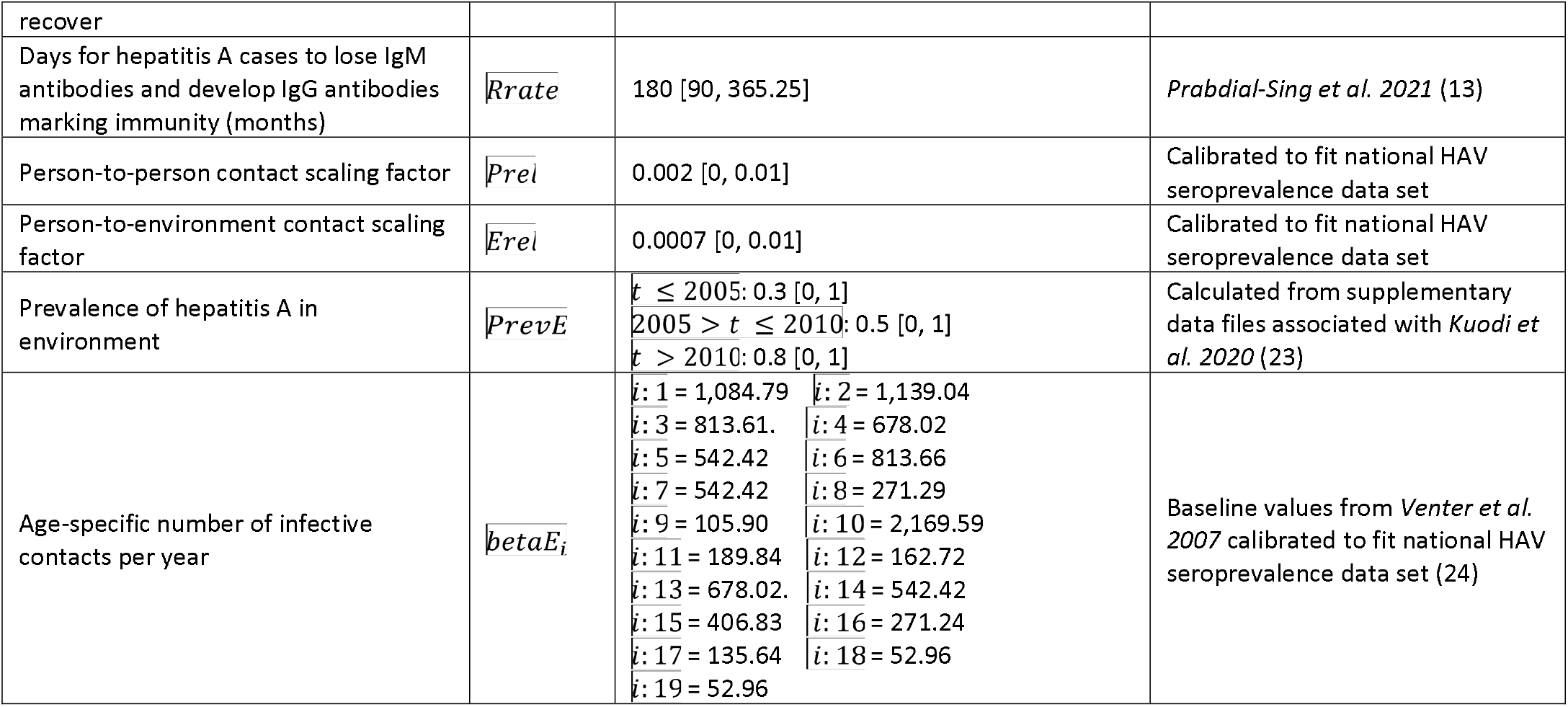
Parameter values and distributions

**Table 5.3:**
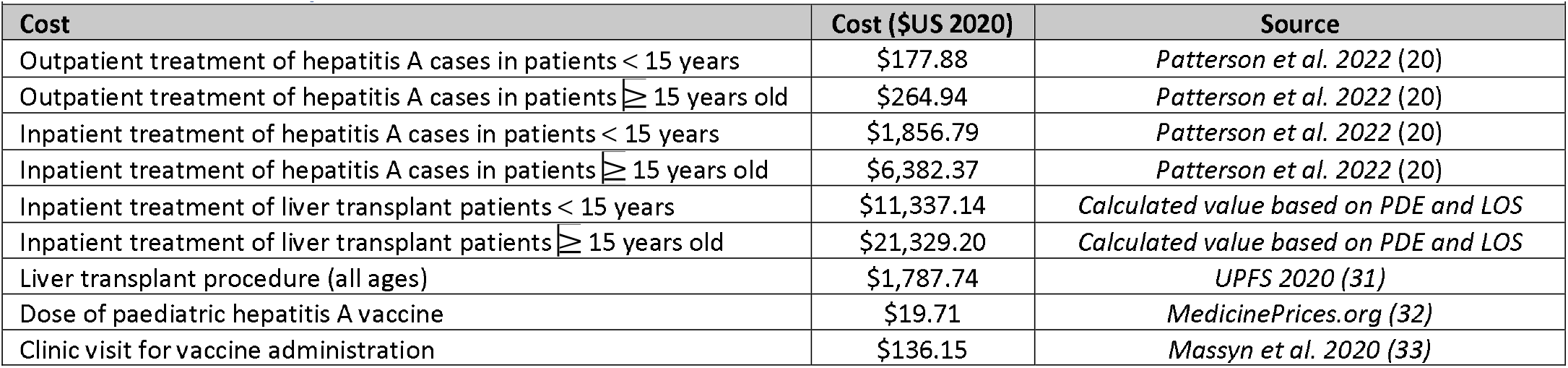
Cost inputs

**Table 5.4:**
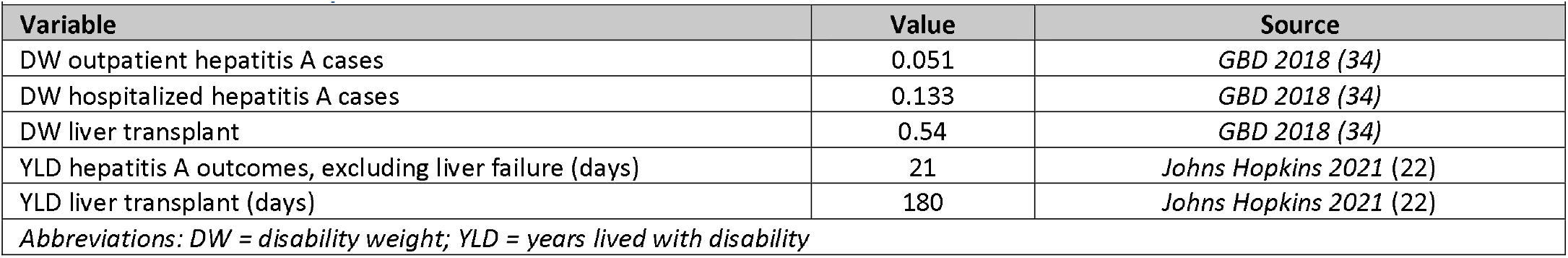
DALY inputs

| Variable | Value | Source |
| --- | --- | --- |
| DW outpatient hepatitis A cases | 0.051 | *GBD 2018 (34)* |
| DW hospitalized hepatitis A cases | 0.133 | *GBD 2018 (34)* |
| DW liver transplant | 0.54 | *GBD 2018 (34)* |
| YLD hepatitis A outcomes, excluding liver failure (days) | 21 | *Johns Hopkins 2021 (22)* |
| YLD liver transplant (days) | 180 | *Johns Hopkins 2021 (22)* |

*Abbreviations: DW = disability weight; YLD = years lived with disability*

**Table 5.5:** Cost-effectiveness of modelled scenarios referencing across a common baseline (2023-2030)

|  |  |  |  |  |  |
| --- | --- | --- | --- | --- | --- |
| Scenario | Total Costs | Incremental Costs | Total DALYs | DALYs averted | Incr. Cost per DALY averted |
| Baseline | \$1,530,392,760 | --- | 27,137 | --- | --- |
| 1 | \$1,714,015,277 | \$183,622,517 | 18,396 | 8,741 | \$21,007 |
| 2 | \$2,009,207,209 | \$478,814,449 | 18,266 | 8,871 | \$53,975 |
| 3 | \$2,195,073,864 | \$664,681,104 | 18,440 | 8,697 | \$76,426 |
| 4 | \$2,851,373,642 | \$1,320,980,882 | 19,151 | 7,986 | \$165,412 |
| *The Incremental costs and DALYs averted presented in this table are calculated by referencing across the common baseline.* |  |  |  |  |  |
| *Abbreviations: Incr. = incremental; DALYs = Disability adjusted life years* |  |  |  |  |  |

Births are classified according to the presence of maternal antibodies (*propM*) into the ***M*** (maternal antibody) and ***S*** (susceptible) compartments. Hepatitis A infection occurs in the ***E*** compartment with the age-specific force of infection given by 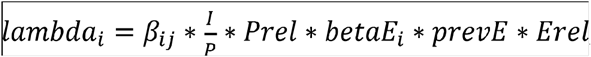, where infection is determined by the number of contacts, the proportion of infected contacts, the transmission probability per contact, the environmental presence of HAV, and the nature of mixing between age groups. The contact pattern between age groups is determined by the conditional probability contact matrix *B*_*ij*_ for South Africa adapted from *Prem et al. 2017* (**Supplementary Table S5.2)** (11).

The ***A*** (asymptomatic) and ***S*** (symptomatic) compartments represents active hepatitis A infections with anti-HAV IgM antibodies following an incubation period *nu*. ***O*** and ***H***_***i***_ represent the treatment sought for uncomplicated hepatitis A cases, while the ***ALF*** compartment represents the treatment sought for viral-induced acute liver failure. Acute liver failure cases spontaneously recover from liver injury into compartment ***ALF***_***R***_, indicate the need for liver transplant and move into compartment ***ALF***_***T***_, or die due to liver injury without transplant in compartment ***ALF***_***D***_. Liver transplant cases recover in compartment ***T***_***R***_ at rate *gammaT* or die following the transplant procedure in compartment ***T***_***D***_ at rate *TDrate*. Hospitalized and outpatient cases lose infectivity at the rate of *gamma* and move into the ***N*** compartment representing previous hepatitis A cases with anti-HAV IgG antibodies that may still have present anti-HAV IgM antibodies. ***R*** represents fully recovered hepatitis A cases with anti-HAV IgG antibodies and no anti-HAV IgM antibodies, while ***D*** represents all death due to hepatitis A infection.

### 1.3.2 Model calibration

The model is fitted to annual South African hepatitis A seroprevalence (anti-HAV IgG) data between 2005 to 2015 from the National Institute of Communicable Diseases (NICD) (12, 13). Ethical approval for the use of this data was obtained from the University of Cape Town Human Research Ethics Committee and the National Institute of Communicable Diseases (NICD). The observed rising trend in hepatitis A seroprevalence data suggests an increase in the incidence of hepatitis A infections (anti-HAV IgM) in South Africa across all age groups. The increase in hepatitis A seroprevalence, however, is not enough to reach the definition of high HAV endemicity as seroprevalence remains <90% for children and adolescents <15 years old between 2005-2015.

The model was run from 2000 with parameters in **Table 5.2** to reach a steady state before being fitted through maximum likelihood estimation to the seroprevalence data from 2005 to 2015. The incidence of HAV seroprevalence in 2015 was considered as baseline for future predictions and all parameters from 2015 were held constant for scenario testing. The NICD seroprevalence data and model seroprevalence outputs are compared by age group in **Figure 5.2**.

Owing to uncertainty in the dataset and a large number of unknown parameters, a simulation approach was selected for data fitting We simulated 100,000 Latin Hypercube Sampled parameter combinations to calibrate the model to key features in the dataset. As the South African testing volumes, IgM positivity rates, and age specific anti-HAV seroprevalence rates varied by year, the model was calibrated to three conditions (features) estimated from the NICD seroprevalence data. As the volume of anti-HAV total antibody tests and proportion of positive total antibody results was highest in 2011, this was chosen as the most reliable year of reporting (12). Only those parameter sets from model runs that reproduced the following criteria were deemed suitable for further analysis:

- seroprevalence below 90% for individuals <20 years old between 2005-2015; and
- seroprevalence to only reach ≥90% in individuals 20-29 years old in 2011 and 2012; and
- seroprevalence below 60% for individuals <5 years old after 2012.

We accepted 1,513 of the 100,000 parameter combinations used to simulate the model reproduced the epidemiological criteria above. The calibration negative log likelihood results are displayed in **Figure 5.3**.

### 1.3.3 Scenario analyses

We used the calibrated model with accepted parameter sets to evaluate various hepatitis A vaccination scenarios from 2023 to 2030. We assessed how each scenario impacted the number of symptomatic hepatitis A cases, hepatitis A mortality, total costs, and total DALYs as compared to the baseline of no vaccination until 2030. The median values are reported for all model outcomes with associated interquartile ranges. In each scenario, the administration of vaccine doses 1 and 2 began in 2023 and catch-up doses began in 2027. The vaccination coverage rates were assumed to be equal to average performance estimates of the EPI-SA in 2019 in relevant age groups and were estimated to be 80%, 60%, and 40% for dose 1, dose 2, and catch-up doses, respectively (25). Vaccine efficacy estimates taken from published literature for dose 1 and subsequent doses were estimated to be 98% and 95%, respectively (26).

**Baseline Scenario:** No vaccination

**Scenario 1:** Dose 1 administered in children < 2 years old

**Scenario 2:** Dose 1 administered in children < 2 years old + Dose 2 administered in children < 3 years old

**Scenario 3:** Dose 1 administered in children < 2 years old + Dose 2 administered in children < 3 years old + Catch-up dose administered in children < 5 years old

**Scenario 4:** Dose 1 administered in children < 2 years old + Dose 2 administered in children < 3 years old + Catch-up dose administered in children < 10 years old

### 1.3.4 Estimation of hepatitis A treatment and routine immunization costs

We conducted the economic evaluation in accordance with CHEERS Guidelines (27). We adopted a provider’s perspective that requires the inclusion of direct health care costs to estimate the cost-effectiveness of the scenarios. The direct costs included treatment costs of HAV and the costs of vaccination. Treatment costs included costs for outpatient care, hospitalization, and liver transplants. Cost inputs displayed in **Table 5.3** were taken from published literature. Where costs were reported in South African Rands (ZAR), they were adjusted to ZAR 2020 using the South African medical consumer price index (CPI) and converted to 2020 United States Dollars (USD) using an average exchange rate over 2020 (US$ 1= ZAR$ 16.61) (28, 29). Where costs were reported in USD, they were converted to ZAR using the relevant exchange rate and adjusted to ZAR 2020 using the South African medical CPI, and then converted back to USD using the 2020 exchange rate.

The cost inputs displayed in **Table 5.3** for hepatitis A outpatient and inpatient treatment at tertiary healthcare facilities were taken from *Patterson et al. 2022* (20). The cost of liver transplant was broken down into treatment of transplant cases and cost of transplant procedures at tertiary healthcare facilities. The cost of treatment for liver transplant cases was calculated by multiplying the cost per inpatient day equivalent (PDE) ($539.86 for patients < 15 years and $821.12 for patients ≥15 years old) by the average length of stay (LOS) (26 days) (20, 30). The cost of liver transplant was taken from the Department of Health Uniform Patient Fee Schedule (UFPS) 2020 to include the procedure and specialist practitioner fee for liver transplants at public tertiary facilities (31). We applied an access parameter of 30% to the cost of liver transplant as not all patients who indicate the need for liver transplant in South Africa will receive one due to social contraindications. To qualify for a transplant, social and socioeconomic criteria are used as exclusion criteria for patients as transplant requires adherence to lifelong treatment and the presence of social support structures for positive outcomes.

Vaccination cost inputs were comprised of the cost per vaccine dose and cost of vaccine administration (clinic visit). The mean cost per vaccine dose was calculated as the average of the single exit prices reported for *Havrix junior single dose vial 0*.*5ml* and *Avaxim prefilled syringe 80 0*.*5ml* (32). As the vaccination scenarios modelled did not include the administration combined with vaccines in the EPI, the cost per vaccine clinic visit was sourced from the District Health Barometer 2020 Public Health Clinic (PHC) expenditure and added to the cost per dose (33).

We calculated disability-adjusted life-years (DALYs) by applying a disability weight (DW) to the estimated years lived with disability (YLD) associated with health states in **Table 5.4**. We assumed a disability weight of 0.051 (95% CI 0.032, 0.074) for all outpatient hepatitis A cases based on the Global Burden of Disease Study 2017 disability weigh estimate for moderate acute hepatitis A (34). We assumed a disability weight of 0.133 (95% CI 0.008, 0.190) for all hospitalized patients based on the Global Burden of Disease Study 2017 disability weigh estimate for severe acute hepatitis A (34). We assumed a disability weight of 0.54 from all patients with liver transplant based on the Global Burden of Disease Study 2017 disability weight estimate for terminal phase of liver cancer due to hepatitis B infection (34). Costs and outcomes were discounted at 5% as recommended by the Health Technology Assessment (HTA) guidelines in South Africa (35).

The results of the economic evaluation for each scenario are reported as incremental cost-effectiveness ratios (ICERs) calculated by comparing each scenario to the baseline. The cost-effectiveness of scenarios was judged against the South African cost-effectiveness threshold (CET) of $3,276 per DALY averted (36). The South African CET reported was reported in 2015 and adjusted to ZAR 2020 using the South African medical CPI and then converted to USD using the 2020 exchange rate.

### 1.3.5 Sensitivity analyses

We ran several one-way sensitivity analyses on key cost and DALY parameters for the most desirable vaccination scenario. We conducted sensitivity analyses on the baseline scenario to determine how the total costs of the scenario would vary for the below changes in cost assumptions and discount rates and display the results in a tornado diagram.

- Remove costs of clinic visit for vaccine administration ($136.15)
- Vary the access to liver transplant procedures to 0% and 100%
- Vary the discount rate between 0% and 10%

## 1.4 Results

### 1.4.1 Baseline scenario

Without implementation of any hepatitis A vaccination strategy from 2023, hepatitis A seroprevalence (anti-HAV IgG) in children < 10 years old is estimated to reach 95.87% [IQR: 93.42%-96.11%] by 2030. However, even with this increase in HAV seroprevalence among children < 10 years old, our model projects that the annual number of symptomatic hepatitis A cases is expected to decline by less than 2% from an expected 49,778 [IQR: 31,546, 87,872] symptomatic case in 2023 to 48,878 [31,057, 87,067] symptomatic cases in 2030. In addition, our model projects that annual hepatitis A mortality will decline by less than 4% from an expected 11,924 [IQR: 8,621-16,446] deaths due to hepatitis A in 2023 to 11,536 [IQR: 8,342, 16,076] deaths in 2030.

**Figure 5.4** shows the impact of each vaccination scenario on symptomatic hepatitis A cases and mortality over the period of 2023-2030.

**Scenario 1:** Administration of one dose of the hepatitis A vaccine in children < 2 years old requires approximately 5.3 million vaccine doses over 2023-2030. The model projects Scenario 1 would avert a total of 136,042 symptomatic cases [IQR: 88,842-221,483] and 31,106 [IQR: 22,975-36,742] deaths due to hepatitis A over the period of 2023 to 2030. Under Scenario 1, one symptomatic case would be averted for approximately every 39 vaccines administered. Similarly, one death due to hepatitis A would be averted for approximately every 171 vaccines administered.

**Scenario 2:** Administration of a first dose of the hepatitis A vaccine in children < 2 years old and a second dose in children < 3 years old requires approximately 7.8 million vaccine doses over 2023-2030. The model projects Scenario 2 would avert a total of 255,857 [IQR: 159,721-225,065] symptomatic cases and 31,585 [IQR: 23,388-37,240] deaths due to hepatitis A over the period of 2023 to 2030. Under Scenario 2, one symptomatic case would be averted for approximately every 56 vaccines administered. Similarly, one death due to hepatitis A would be averted for approximately every 247 vaccines administered.

**Scenario 3:** Administration of a first dose of the hepatitis A vaccine in children < 2 years old and a second dose in children < 3 years old with a catch-up dose administered to children < 5 years old that are not already vaccinated requires approximately 9.2 million vaccine doses over 2023-2030. The model projects that Scenario 3 would avert a total of 259,318 [IQR: 162,828-477,574] symptomatic cases and 30,982 [IQR: 22,502-37,488] deaths due to hepatitis A over the period of 2023 to 2030. Under Scenario 3, one symptomatic case would be averted for approximately every 68 vaccines administered. Similarly, one death due to hepatitis A would be averted for approximately every 298 vaccines administered.

**Scenario 4:** Administration of a first dose of the hepatitis A vaccine in children < 2 years old and a second dose in children < 3 years old with a catch-up dose administered to children < 10 years old not already vaccinated requires approximately 11.7 million vaccine doses over 2023-2030. The model projects that Scenario 4 would avert a total of 267,947 [IQR: 169,625-482,796] symptomatic cases and 29,890 [IQR: 21,235-37,309] deaths due to hepatitis A over the period of 2023 to 2030. Under Scenario 4, one symptomatic case would be averted for approximately every 86 vaccines administered. Similarly, one death due to hepatitis A would be averted for approximately every 392 vaccines administered.

### 1.4.2 Cost-effectiveness of vaccination

For the modelled scenarios, the median estimated cost of the different vaccination strategies ranged from $1.71 billion to $2.85 billion over the period of 2023 to 2030, with the cost increasing for each successive scenario and approximately 39-52% of costs being due to vaccination. The ICERs for the vaccination scenarios in **Table 5.3** were calculated by comparing each scenario to the baseline. In **Supplementary Table S5.3**, we also present ICERS calculated by comparing each scenario to the previous undominated and less costly scenario. The cost-effectiveness of scenarios was judged against the South African CET of $3,276 per DALY averted (36).

The model suggests that implementation of all potential vaccination scenarios would deliver health gains in the population, with the lowest incremental cost per DALY averted against baseline for Scenario 1. The model projects that Scenario 1, representing administration of a single dose of hepatitis A vaccine in children < 2 years old from 2023 to 2030, would avert 8,741 DALYs, however is not cost-effective against the CET with an ICER per DALY averted of $21,006. In **Supplementary Table S5.3**, the results of our model show that Scenarios 3 and 4 were absolutely dominated in that they produced less health gains and were more expensive than Scenarios 1 and 2. These results signal that the timing of vaccination is critical in the roll-out of potential hepatitis A prevention programs. While Scenario 3 and 4 include the administration of more vaccine doses and avert more symptomatic cases of hepatitis A, the total health gains are smaller than in Scenarios 1 and 2 owing to the population being infected before vaccination through the mass campaigns at older ages. With our results, the model suggests that natural exposure to HAV may begin as early as 3 years old in South Africa.

### 1.4.3 Sensitivity analysis

Our one-way sensitivity analysis 4 on the total cost of Scenario 1 reported in **Figure 5.5** shows that varying access to liver transplant between 0% and 100% has the largest impact in results (total cost delta = $609,302,599). When increasing the access to liver transplant to 100% for baseline and Scenario 1, the ICER for Scenario 1 becomes cost-effective against the CET (ICER = $2,425) **(Supplementary Table S5.4)**.

## 1.5 Discussion

Our results indicate that administration of a single dose of the hepatitis A vaccine in children < 2 years old in South Africa between the period of 2023 to 2030 would produce significant health gains. The implementation of this vaccination strategy between 2023 and 2030 has the potential to avert a total of 136,042 symptomatic cases [IQR: 88,842-221,483] and 31,106 [IQR: 22,975-36,742] deaths due to hepatitis A. The model projects that for every 39 hepatitis A vaccines administered, one symptomatic case of hepatitis A would be averted. Similarly, for every 171 hepatitis A vaccines administered, one death due to hepatitis A would be averted. Our results show that the implementation of a single dose of the hepatitis A vaccine in children < 2 years old in South Africa would avert 8,741 DALYs over the period of 2023-2030, however is not cost-effective against the South African CET with an ICER per DALY averted of $21,006.

The total cost of implementing a single dose of the hepatitis A vaccine for children < 2 years old over the eight-year intervention period is estimated to be $1.71 billion, with approximately 39% of the cost due to the 5.3 million vaccine doses required. When reviewing the total cost of modelled scenarios, it is notable that less than 50% of the total costs were due to vaccination. These results indicate that the burden of hepatitis A in the baseline scenario is heavy for the healthcare system and national health budget in South Africa.

Our study signals that the timing of hepatitis A vaccine administration is important as Scenarios 3 and 4 were absolutely dominated by Scenarios 1 and 2. While Scenario 3 and 4 include the administration of more vaccine doses and avert more symptomatic cases of hepatitis A, the total health gains are less than in Scenarios 1 and 2 owing to the population being infected before vaccination through the mass campaigns at older ages.

In regard to patient outcomes, we applied a liver transplant access parameter of 30% in our economic evaluation as not all patients who indicate the need for liver transplant in South Africa will receive one due to social contraindications. To qualify for a transplant, social and socioeconomic criteria are used as exclusion criteria for patients as transplant requires adherence to lifelong treatment and the presence of social support structures for positive outcomes. Our sensitivity analysis shows that the cost-effectiveness of vaccination was highly sensitivity to varying access to liver transplant. When increasing the access to liver transplant to 100% for baseline and Scenario 1, the ICER for Scenario 1 becomes cost-effective against the CET (ICER = $2,425). Given these findings, we recommend further research is conducted to understand the access to liver transplants in South Africa to better estimate the cost of liver transplant care for hepatitis A patients and cost-effectiveness of vaccination.

The main strength of this study is that, to the best of our knowledge, it is the first to utilize a dynamic modelling approach to understand the epidemiology of hepatitis A in South Africa and to conduct a cost-effectiveness analysis of routine hepatitis A vaccination in the country. Our study uses local cost data drawn from a retrospective folder review of hepatitis A cases requiring outpatient care or hospitalization in South Africa and this contextually relevant data leads to the derivation of more realistic cost projections in the country.

The modelling presented in this paper has been used to develop a <u>user-friendly application</u> for vaccine policy makers to further interrogate the model outcomes and consider the costs and benefits of introducing routine hepatitis A vaccination in South Africa. The application allows users to vary clinical parameters in the model such as the proportion of hepatitis A patients that require hospitalisation or develop viral-induced liver failure as well as associated costs. Once the user has varied these parameters, they have the opportunity to develop vaccination programs and compare outcomes to assess the potential cost-effectiveness. The application has been developed in R using the Rshiny package and can be accessed using this link (https://masha-app.shinyapps.io/HepA-VacExplorer/).

Several limitations must be considered in the interpretation of our results from the hepatitis A transmission model. It is important to take into account that incidence rates for hepatitis A are likely underreported due to the circumstances and mild nature with which the disease can present. In addition, the transmission model assumes that all symptomatic cases seek treatment for infection, which may not be the case. As these estimates were missing from the literature, we recommend more research be conducted on treatment seeking behaviours for patients with hepatitis A.

It should also be noted that the projected increase in hepatitis A seroprevalence among children < 10 years old in South Africa is unexpected and these results should be interpreted with caution. While the model was calibrated using the largest description of HAV seroprevalence within South Africa to date, the HAV seroprevalence data published by the NICD was unable to determine yearly seroprevalence trends due to the low volumes of anti-HAV total antibody testing and uneven distribution among age groups (12). The data that we used to calibrate the model was available only until 2015, which means caution should be applied when interpreting forecasted results until 2030. In addition, we were unable to determine a trend in the environmental presence of HAV which plays a large part in childhood hepatitis A transmission. To validate and update the model’s seroprevalence projections, new data on anti-HAV IgG and IgM positivity and the environmental presence of HAV in South Africa should be included in the model as it comes available. Further analysis should include fitting the model to a decreasing trend in HAV seroprevalence between 2005 and 2015. Other limitations of this study include that the cost of hepatitis A inpatient treatment is likely overestimated as it is drawn from a tertiary hospital setting.

## 1.6 Conclusion

The results of this study indicate that implementation of a single dose of the hepatitis A vaccine in South African children < 2 years old between 2023 and 2030 generates health gains in comparison to the baseline approach, however, is not cost-effective against the CET with an ICER per DALY averted of $21,006. Given the sensitivity of the model to varying access to liver transplant, we recommend further research is conducted to understand the access parameters in order to better inform considerations of hepatitis A vaccination policies. In addition, further analysis using this model might include fitting the model to a decreasing trend in HAV seroprevalence between 2005 and 2015.

## Data Availability

All data produced in the present study are available upon reasonable request to the authors.

## 1.8 Acknowledgements

Many thanks to Nishi Prabdial-Sing for providing raw IgM data from the publication “*Establishment of Outbreak Thresholds for Hepatitis A in South Africa Using Laboratory Surveillance, 2017–2020*”.

