## Supplementary Files for "Modelling the cost-effectiveness of hepatitis A vaccination in South Africa"

### 1 Appendices

#### 1.1 Ethics approval documents

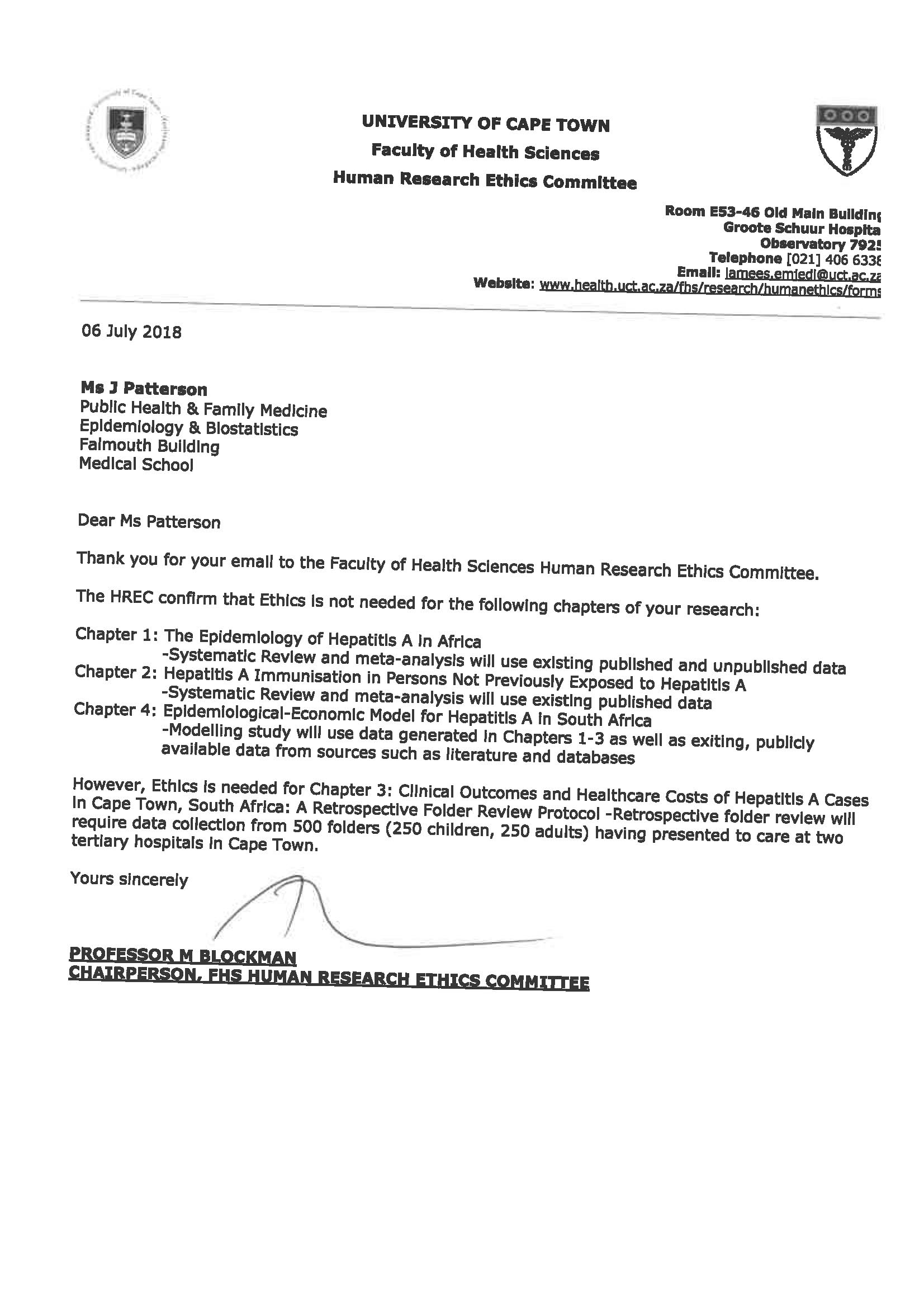

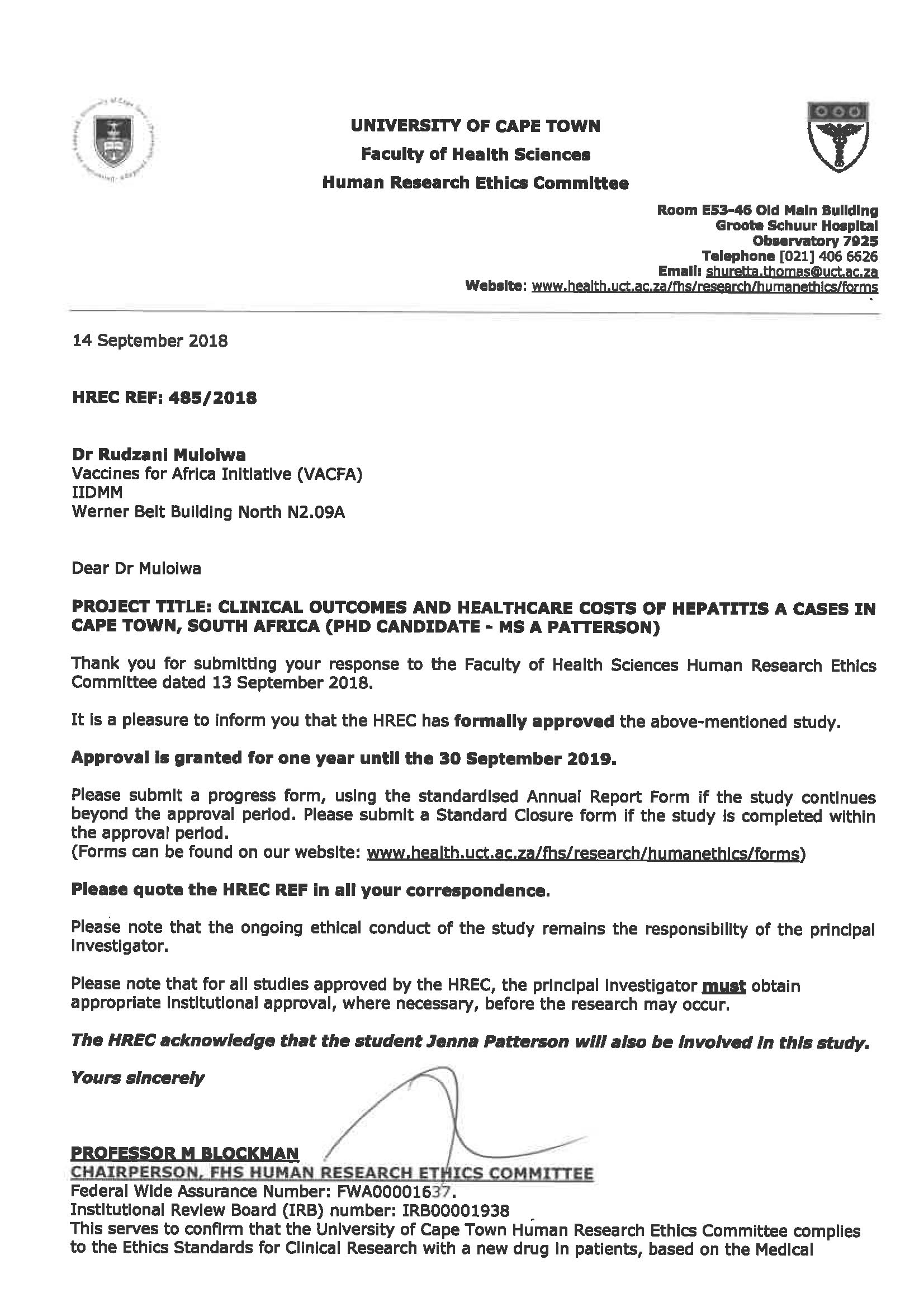

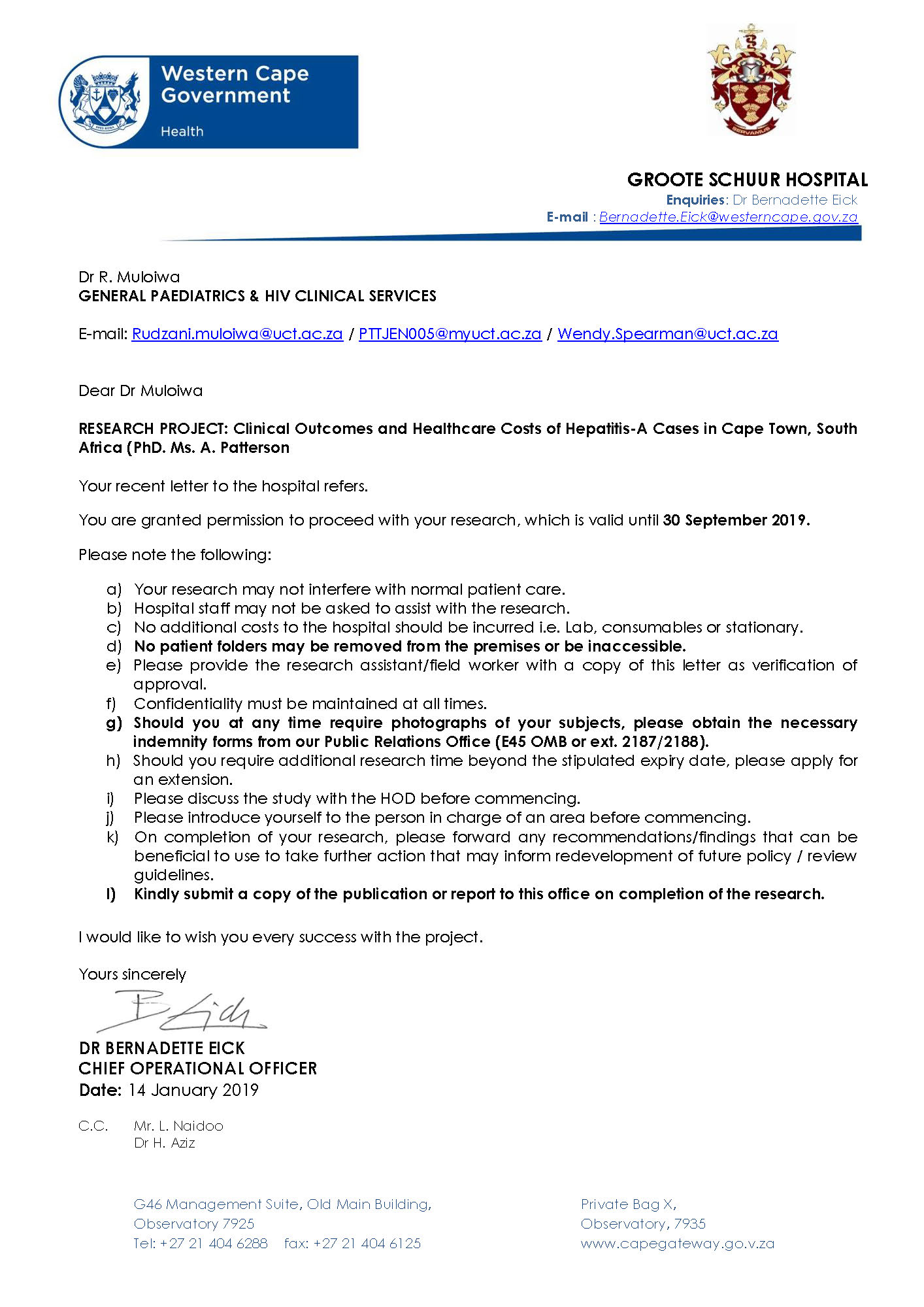

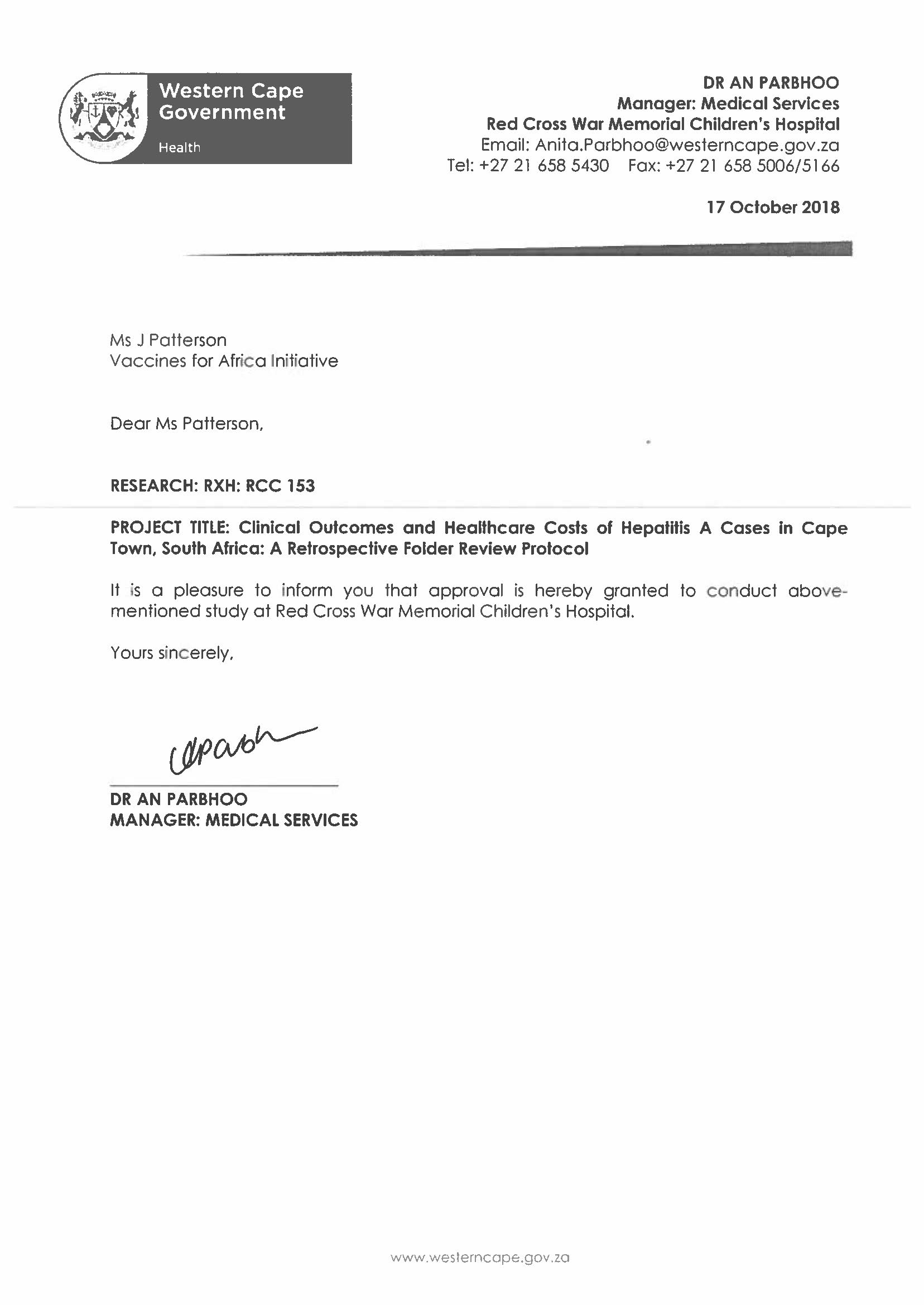

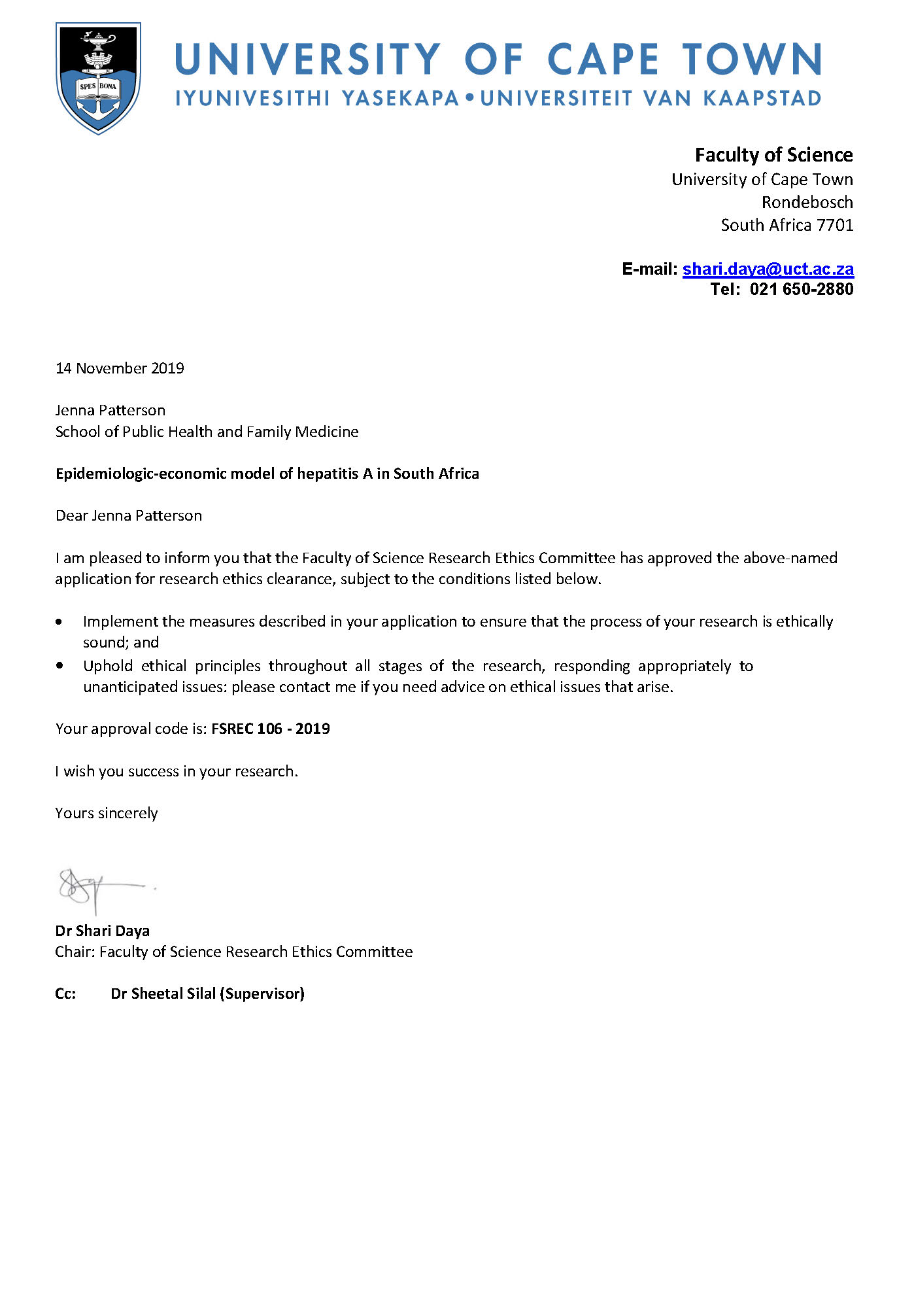

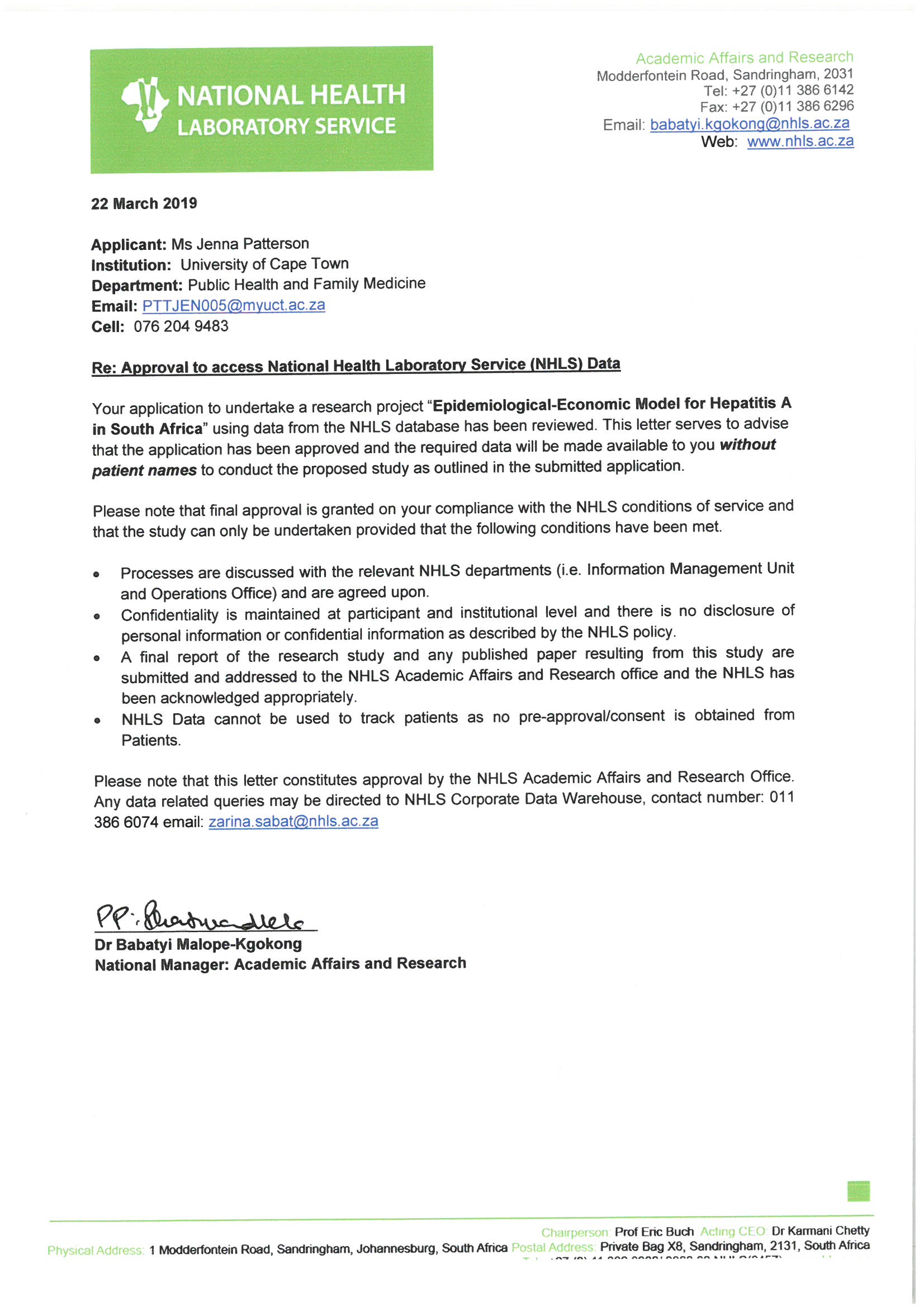

#### 1.2 Supplementary tables

| Supplementary Table S2.1 - GRADE Table | | | |
| --- | --- | --- | --- |
| **Outcome: Epidemiological transition from high to intermediate or low HAV endemicity in South Africa** | | | |
|  | **Rating** | **Footnotes** | **Quality of evidence** |
| **Study design** | High | Systematic review | Moderate |
| **Risk of bias** | Low | Minimal risk of bias among included studies |  |
| **Inconsistency** | Low | Minimal inconsistency for South African data |  |
| **Indirectness** | Very low | Not detected |  |
| **Imprecision** | Very low | Not detected |  |
| **Publication bias** | Very low | Not detected |  |

| Supplementary Table S3.1: Risk of bias judgements for included studies | | | | | | | | | | | |
| --- | --- | --- | --- | --- | --- | --- | --- | --- | --- | --- | --- |
| **Study ID** | **Representation of the national population** | **Representation of target population** | **Random selection or census** | **Minimal likelihood of non-response bias** | **Data collected directly from participants** | **Acceptable case definition** | **Valid measurement** | **Same mode of data collection** | **Appropriate length** | **Appropriate numerator(s) and denominator(s)** | **Score** |
| Alam et al., 2009 | No | Yes | No | Yes | Yes | Yes | Yes | Yes | Yes | Yes | 8 |
| Asim et al., 2009 | Yes | Yes | No | Yes | Yes | Yes | Yes | Yes | Yes | Yes | 9 |
| Bechmann et al., 2014 | Yes | Yes | Yes | Yes | Yes | Yes | Yes | Yes | Yes | Yes | 10 |
| Bhati et al., 2013 | Yes | No | No | Yes | Yes | Yes | Yes | Yes | Yes | Yes | 8 |
| Borkakoti et al., 2013 | No | Yes | No | Yes | Yes | Yes | Yes | Yes | Yes | Yes | 8 |
| Bravo et al., 2012 | No | No | Yes | Yes | Yes | Yes | Yes | Yes | Yes | Yes | 8 |
| Cervio et al., 2011 | No | Yes | Yes | Yes | Yes | Yes | Yes | Yes | Yes | Yes | 9 |
| Das et al., 2016 | Yes | Yes | No | Yes | Yes | Yes | Yes | Yes | Yes | Yes | 9 |
| Gupta et al., 2015 | Yes | Yes | Yes | Yes | Yes | Yes | Yes | Yes | Yes | Yes | 10 |
| Ho et al., 2014 | Yes | Yes | Yes | Yes | Yes | Yes | Yes | Yes | Yes | Yes | 10 |
| Latif et al., 2010 | No | Yes | No | Yes | Yes | Yes | Yes | Yes | Yes | Yes | 9 |
| Mamun et al., 2009 | No | Yes | No | No | Yes | Yes | Yes | Yes | Yes | Yes | 9 |
| Manka et al., 2015 | No | Yes | Yes | No | Yes | Yes | No | Yes | Yes | Yes | 8 |
| Mendizabal et al., 2014 | No | Yes | Yes | No | Yes | Yes | Yes | Yes | Yes | Yes | 8 |
| Mishra et al., 2016 | No | Yes | No | No | Yes | Yes | Yes | Yes | Yes | Yes | 7 |
| Mumtaz et al., 2009 | Yes | Yes | Yes | Yes | Yes | Yes | Yes | Yes | Yes | Yes | 10 |
| Pandit et al., 2015 | No | Yes | No | No | Yes | Yes | Yes | Yes | Yes | Yes | 8 |
| Poovorawan et al., 2013 | Yes | Yes | Yes | Yes | Yes | Yes | Yes | Yes | Yes | No | 9 |
| Schwarz et al., 2014 | Yes | Yes | Yes | Yes | Yes | Yes | Yes | Yes | Yes | No | 9 |
| Shalimar et al., 2017 | Yes | Yes | Yes | Yes | Yes | Yes | Yes | Yes | Yes | Yes | 10 |
| Silverio et al., 2015 | Yes | Yes | Yes | Yes | Yes | Yes | Yes | Yes | Yes | Yes | 10 |
| Somasekar et al., 2017 | Yes | Yes | No | No | Yes | Yes | Yes | Yes | Yes | Yes | 8 |
| Uddin Jamro et al., 2013 | Yes | Yes | Yes | Yes | Yes | Yes | Yes | Yes | Yes | Yes | 10 |
| Tsunoda et al., 2017 | Yes | Yes | Yes | No | Yes | Yes | Yes | Yes | Yes | Yes | 9 |
| Zhao et al., 2014 | Yes | Yes | No | No | Yes | Yes | Yes | Yes | Yes | Yes | 8 |

| Supplementary Table S4.1: Mean and median lengths of hospitalization by patient outcome | | |
| --- | --- | --- |
| **Adult patients** | | |
| **Patient Outcome** | **Mean length of stay in days (95% CI)** | **Median length of stay in days (IQR)** |
| Uncomplicated (n=180) | 5.0 (3.9, 6.1) | 0.8 (0.3, 2.1) |
| Complicated (n=29) | 14.2 (0.7, 27.8) | 4.1 (2.0, 10.7) |
| Deceased (n=3) | 1.8 (1.8, 1.8) | 1.8 (1.8, 1.8) |
| **Paediatric patients** | | |
| **Patient Outcome** | **Mean length of stay in days (95% CI)** | **Median length of stay in days (IQR)** |
| Uncomplicated (n=211) | 1.3 (1.1, 1.6) | 0.3 (0.2, 0.9) |
| Complicated (n=27) | 8.1 (3.5, 12.8) | 5.4 (1.5, 7.3) |
| Deceased (n=1) | 5.3 (5.3, 5.3) | 5.5 (5.5, 5.5) |

| Supplementary Table S4.2: Unit counts and cost (2018 USD) for patient-specific hepatitis A items | | | | | |
| --- | --- | --- | --- | --- | --- |
| **Blood tests** | | | | | |
| **Test** | **Unit counts for adult hepatitis A patient population** | **Unit counts for paediatric hepatitis A patient population** | **Unit cost in USD** | **Total cost USD for adult hepatitis A patient population** | **Total cost USD for paediatric hepatitis A patient population** |
| HAV IgG | 9 | 3 | 8.96 | 80.65 | 26.88 |
| HAV IgM | 219 | 224 | 8.96 | 1962.54 | 2007.34 |
| HBsAg | 128 | 105 | 8.96 | 1147.05 | 940.94 |
| HBC | 63 | 62 | 8.96 | 564.57 | 555.60 |
| HCV | 94 | 82 | 8.96 | 842.37 | 734.83 |
| ALT | 358 | 288 | 3.23 | 1157.74 | 931.36 |
| AST | 306 | 234 | 3.23 | 989.57 | 756.73 |
| ALP | 309 | 214 | 3.08 | 951.93 | 659.27 |
| Albumin | 201 | 145 | 2.86 | 575.61 | 415.24 |
| Total bilirubin | 325 | 243 | 2.51 | 816.14 | 610.22 |
| Conjugated bilirubin | 275 | 208 | 1.91 | 525.95 | 397.81 |
| GGT | 299 | 206 | 3.23 | 966.94 | 666.18 |
| Fibrinogen | 23 | 41 | 2.43 | 56.00 | 99.82 |
| INR | 273 | 247 | 3.37 | 919.50 | 831.93 |
| HB | 230 | 199 | 1.28 | 295.18 | 255.39 |
| FBC Differential Count | 229 | 192 | 2.26 | 518.55 | 434.77 |
| Full Blood Count Inch Platelet | 222 | 190 | 4.13 | 916.75 | 784.60 |
| Neutrophils | 37 | 43 | 2.26 | 83.78 | 97.37 |
| Na | 197 | 166 | 2.16 | 425.65 | 358.67 |
| K+ | 207 | 167 | 2.16 | 447.26 | 360.83 |
| Urea | 191 | 161 | 2.16 | 412.69 | 347.87 |
| Creatine | 224 | 184 | 2.16 | 483.99 | 397.56 |
| HIV | 57 | 67 | 3.93 | 223.90 | 263.19 |
| **Radiology** | | | | | |
| **Test** | **Unit counts for adult hepatitis A patient population** | **Unit counts for paediatric hepatitis A patient population** | **Unit cost in USD** | **Total cost USD for adult hepatitis A patient population** | **Total cost USD for paediatric hepatitis A patient population** |
| AXR | 5 | 2 | 4.47 | 22.37 | 8.95 |
| Abdominal ultrasound | 50 | 36 | 12.68 | 633.90 | 456.41 |
| Liver ultrasound | 1 | 1 | 7.86 | 7.86 | 7.86 |
| Gastroscopy | 1 | 1 | 74.92 | 74.92 | 74.92 |
| Brain CT | 4 | 0 | 60.75 | 242.98 | 0.00 |
| **Medicines and products** | | | | | |
| **Medicines and products** | **Number of prescriptions in adult hepatitis A patient population** | **Number of prescriptions in paediatric hepatitis A patient population** | **Mean unit cost USD per prescription** | **Total cost USD for prescriptions in adult hepatitis A population** | **Total cost USD for prescriptions in paediatric hepatitis A population** |
| Antibiotics | 27 | 31 | 5.33 | 1524.56 | 3019.70 |
| Antifungals | 11 | 5 | 0.79 | 112.13 | 74.75 |
| Antiemetics | 41 | 2 | 0.97 | 38.56 | 575.28 |
| Lactulose | 14 | 3 | 1.31 | 251.08 | 77.51 |
| Steroid | 2 | 2 | 4.71 | 75.84 | 202.22 |
| Vitamin K | 33 | 24 | 2.97 | 1770.66 | 729.50 |
| Other medicines | 128 | 53 | 0.98 | 1841.09 | 785.13 |
| Fresh frozen plasma | 1 | 0 | 157.79 | 2327.44 | NA |
| Platelets | 2 | 0 | 618.97 | 18247.26 | NA |
| Prescriptions at discharge | 145 | 84 | 13.52 | 2149.74 | 946.64 |

###### Supplementary Table S5.1: Ordinary differential equations

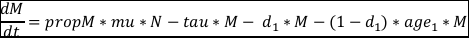

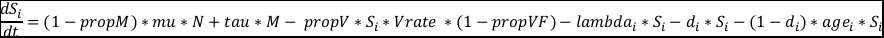

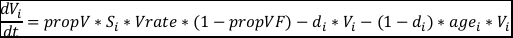

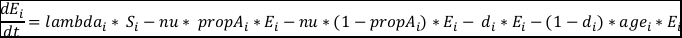

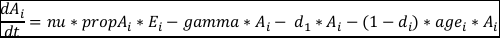

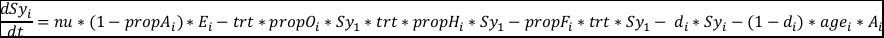

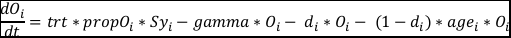

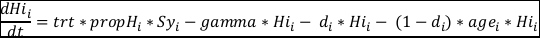

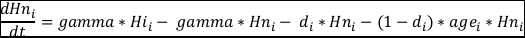

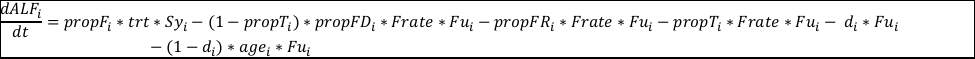

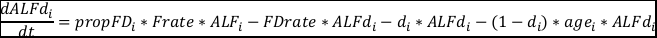

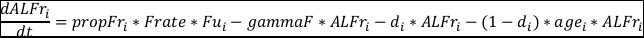

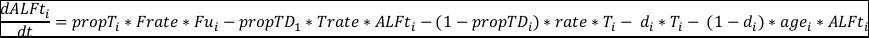

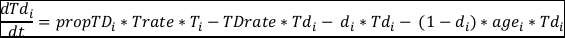

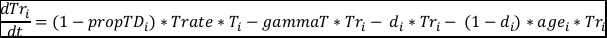

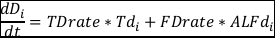

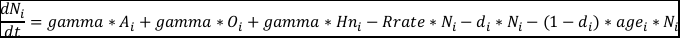

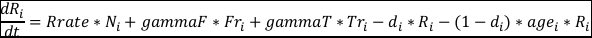

| Supplementary Table S5.2: Daily contact matrix | | | | | | | | | | | | | | | | | | | | |
| --- | --- | --- | --- | --- | --- | --- | --- | --- | --- | --- | --- | --- | --- | --- | --- | --- | --- | --- | --- | --- |
| **Age** | | | | | | | | | | | | | | | | | | | | |
| **Age** |  | 0 | 1 | 2 | 3 | 4 | 5 | 6 | 7 | 8 | 9 | 10 to 14 | 15t o 19 | 20 to 29 | 30 to 39 | 40 to 49 | 50 to 59 | 60 to 69 | 70 to 79 | 80+ |
|  | 0 | 0.13 | 0.13 | 0.13 | 0.12 | 0.12 | 0.13 | 0.13 | 0.13 | 0.12 | 0.12 | 0.17 | 0.11 | 0.46 | 0.52 | 0.20 | 0.13 | 0.06 | 0.02 | 0.00 |
|  | 1 | 0.13 | 0.13 | 0.12 | 0.12 | 0.12 | 0.13 | 0.13 | 0.13 | 0.12 | 0.12 | 0.17 | 0.11 | 0.46 | 0.52 | 0.19 | 0.13 | 0.06 | 0.02 | 0.00 |
|  | 2 | 0.13 | 0.12 | 0.12 | 0.12 | 0.12 | 0.13 | 0.13 | 0.13 | 0.12 | 0.12 | 0.17 | 0.11 | 0.45 | 0.51 | 0.19 | 0.13 | 0.06 | 0.02 | 0.00 |
|  | 3 | 0.12 | 0.12 | 0.12 | 0.12 | 0.12 | 0.12 | 0.12 | 0.12 | 0.12 | 0.12 | 0.17 | 0.11 | 0.45 | 0.51 | 0.19 | 0.13 | 0.06 | 0.02 | 0.00 |
|  | 4 | 0.12 | 0.12 | 0.12 | 0.12 | 0.12 | 0.12 | 0.12 | 0.12 | 0.12 | 0.11 | 0.17 | 0.11 | 0.44 | 0.50 | 0.19 | 0.13 | 0.06 | 0.02 | 0.00 |
|  | 5 | 0.06 | 0.06 | 0.06 | 0.06 | 0.06 | 0.06 | 0.06 | 0.06 | 0.06 | 0.05 | 0.41 | 0.12 | 0.27 | 0.47 | 0.26 | 0.09 | 0.05 | 0.01 | 0.00 |
|  | 6 | 0.06 | 0.06 | 0.06 | 0.06 | 0.06 | 0.06 | 0.06 | 0.06 | 0.06 | 0.06 | 0.41 | 0.12 | 0.28 | 0.48 | 0.26 | 0.09 | 0.05 | 0.01 | 0.00 |
|  | 7 | 0.06 | 0.06 | 0.06 | 0.06 | 0.06 | 0.06 | 0.06 | 0.06 | 0.06 | 0.05 | 0.41 | 0.12 | 0.27 | 0.47 | 0.26 | 0.09 | 0.05 | 0.01 | 0.00 |
|  | 8 | 0.06 | 0.06 | 0.06 | 0.05 | 0.05 | 0.06 | 0.06 | 0.06 | 0.05 | 0.05 | 0.40 | 0.11 | 0.27 | 0.46 | 0.25 | 0.09 | 0.05 | 0.01 | 0.00 |
|  | 9 | 0.05 | 0.05 | 0.05 | 0.05 | 0.05 | 0.05 | 0.06 | 0.05 | 0.05 | 0.05 | 0.38 | 0.11 | 0.26 | 0.44 | 0.24 | 0.09 | 0.05 | 0.01 | 0.00 |
|  | 10 to 14 | 0.12 | 0.12 | 0.12 | 0.11 | 0.11 | 0.12 | 0.12 | 0.12 | 0.11 | 0.11 | 12.55 | 1.31 | 1.22 | 1.60 | 1.46 | 0.42 | 0.17 | 0.07 | 0.02 |
|  | 15 to 19 | 0.07 | 0.07 | 0.07 | 0.07 | 0.07 | 0.07 | 0.07 | 0.07 | 0.07 | 0.07 | 3.74 | 9.68 | 3.30 | 1.56 | 1.75 | 0.58 | 0.16 | 0.04 | 0.01 |
|  | 20 to 29 | 0.13 | 0.13 | 0.13 | 0.13 | 0.12 | 0.13 | 0.13 | 0.13 | 0.12 | 0.12 | 0.95 | 5.04 | 16.20 | 6.20 | 3.84 | 2.04 | 0.53 | 0.08 | 0.02 |
|  | 30 to 39 | 0.24 | 0.24 | 0.23 | 0.23 | 0.23 | 0.24 | 0.24 | 0.24 | 0.23 | 0.22 | 2.37 | 1.38 | 6.41 | 8.32 | 4.64 | 1.84 | 0.68 | 0.10 | 0.02 |
|  | 40 to 49 | 0.23 | 0.22 | 0.22 | 0.22 | 0.21 | 0.22 | 0.23 | 0.22 | 0.22 | 0.21 | 2.25 | 2.50 | 3.75 | 4.94 | 5.00 | 1.79 | 0.53 | 0.10 | 0.02 |
|  | 50 to 59 | 0.20 | 0.20 | 0.20 | 0.19 | 0.19 | 0.20 | 0.20 | 0.20 | 0.19 | 0.19 | 1.93 | 1.89 | 3.57 | 3.18 | 3.11 | 2.19 | 0.67 | 0.11 | 0.03 |
|  | 60 to 69 | 0.14 | 0.14 | 0.14 | 0.14 | 0.13 | 0.14 | 0.14 | 0.14 | 0.14 | 0.13 | 1.11 | 0.84 | 1.92 | 2.61 | 1.84 | 1.33 | 0.96 | 0.18 | 0.02 |
|  | 70 to 79 | 0.08 | 0.08 | 0.08 | 0.08 | 0.08 | 0.08 | 0.09 | 0.08 | 0.08 | 0.08 | 1.20 | 0.95 | 0.65 | 0.98 | 1.13 | 0.77 | 0.56 | 0.34 | 0.09 |
|  | 80+ | 0.10 | 0.09 | 0.09 | 0.09 | 0.09 | 0.10 | 0.10 | 0.09 | 0.09 | 0.09 | 0.60 | 0.48 | 0.26 | 0.41 | 0.51 | 0.38 | 0.19 | 0.12 | 0.04 |

| Supplementary Table S5.3 Cost-effectiveness of modelled scenarios referencing previous undominated approach (2023-2030) | | | | | |
| --- | --- | --- | --- | --- | --- |
| **Scenario** | **Total Costs** | **Incremental Costs** | **Total DALYs** | **DALYs averted** | **Incr. Cost per DALY averted** |
| Baseline | $1,530,392,760 |  | 27,137 |  |  |
| 1 | $1,714,015,277 | $183,622,517 | 18,396 | 8,741 | $21,007 |
| 2 | $2,009,207,209 | $295,191,932 | 18,266 | 130 | $2,270,707 |
| 3 | $2,195,073,864 | $185,866,655 | 18,440 | -174 | ($1,068,199) |
| 4 | $2,851,373,642 | $656,299,778 | 19,151 | -711 | ($923,066) |
| *The Incremental costs and DALYs averted presented in this table are calculated by referencing the previous undominated and less costly scenario.*  *Abbreviations: Incr. = incremental; DALYs = Disability adjusted life years* | | | | | |

| Supplementary Table S5.4: One-way sensitivity analysis for Scenario 1 ICER Results | | | |
| --- | --- | --- | --- |
| **One-way sensitivity analysis** | **Scenario 1 Total Cost** | **DALYS averted against baseline** | **Incr. cost per DALY averted against baseline** |
| Cost of clinic visit removed | $1,128,653,105 | 18,396 | $45,958 |
| Access to liver transplant at 0% | $1,531,224,497 | 18,396 | -$31,048 |
| Access to liver transplant at 100% | $2,140,527,097 | 18,396 | $2,426 |
| Discount rate at 0% | $2,025,301,242 | 20,984 | -$19,972 |
| Discount rate at 10% | $1,477,986,262 | 16,406 | -$22,079 |

#### 1.3 Supplementary figures

###### Supplementary Figure S3.1: Prevalence of hepatitis C virus induced acute liver failure

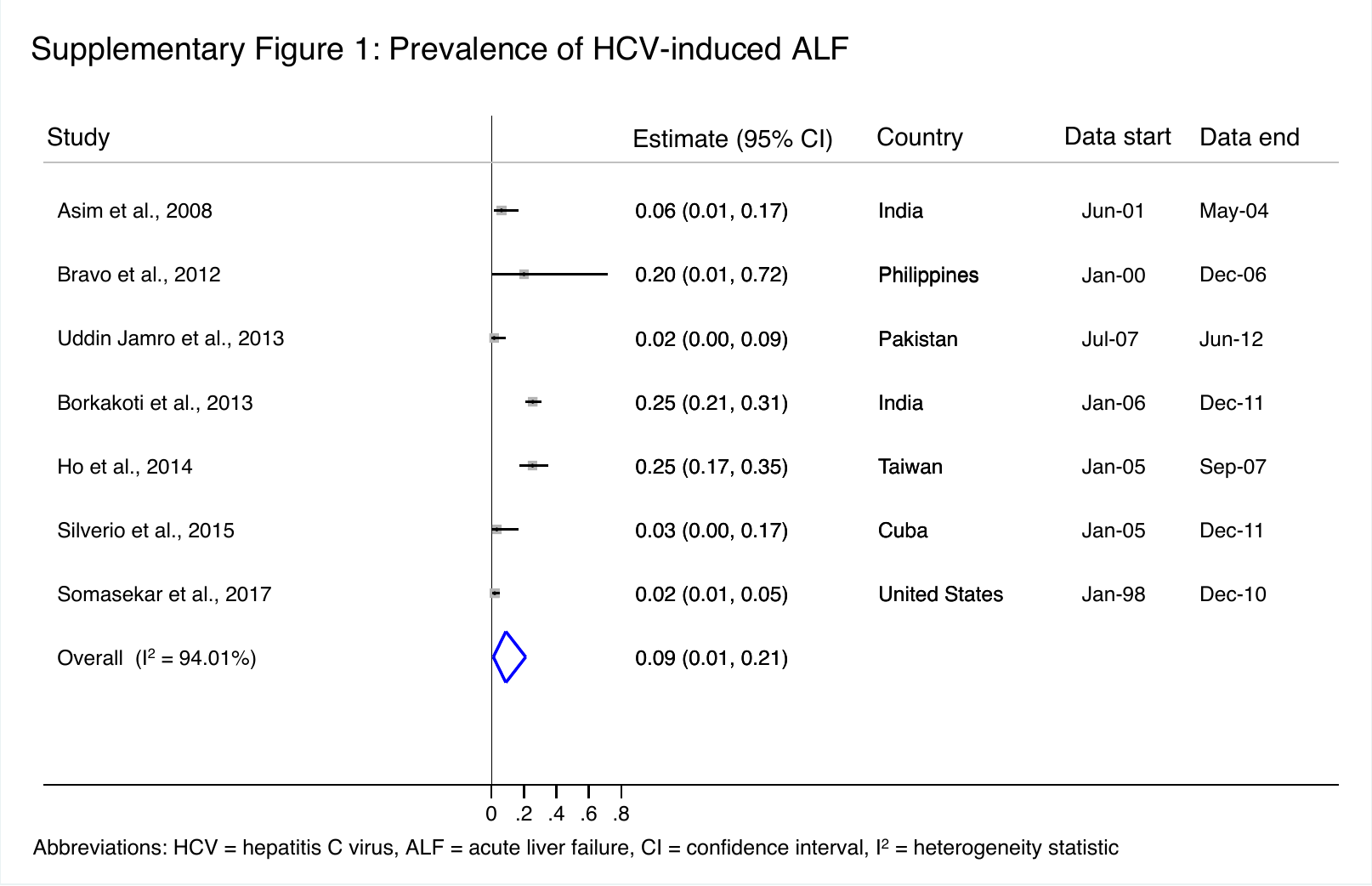

###### Supplementary Figure S3.2: Prevalence of hepatitis E virus induced acute liver failure

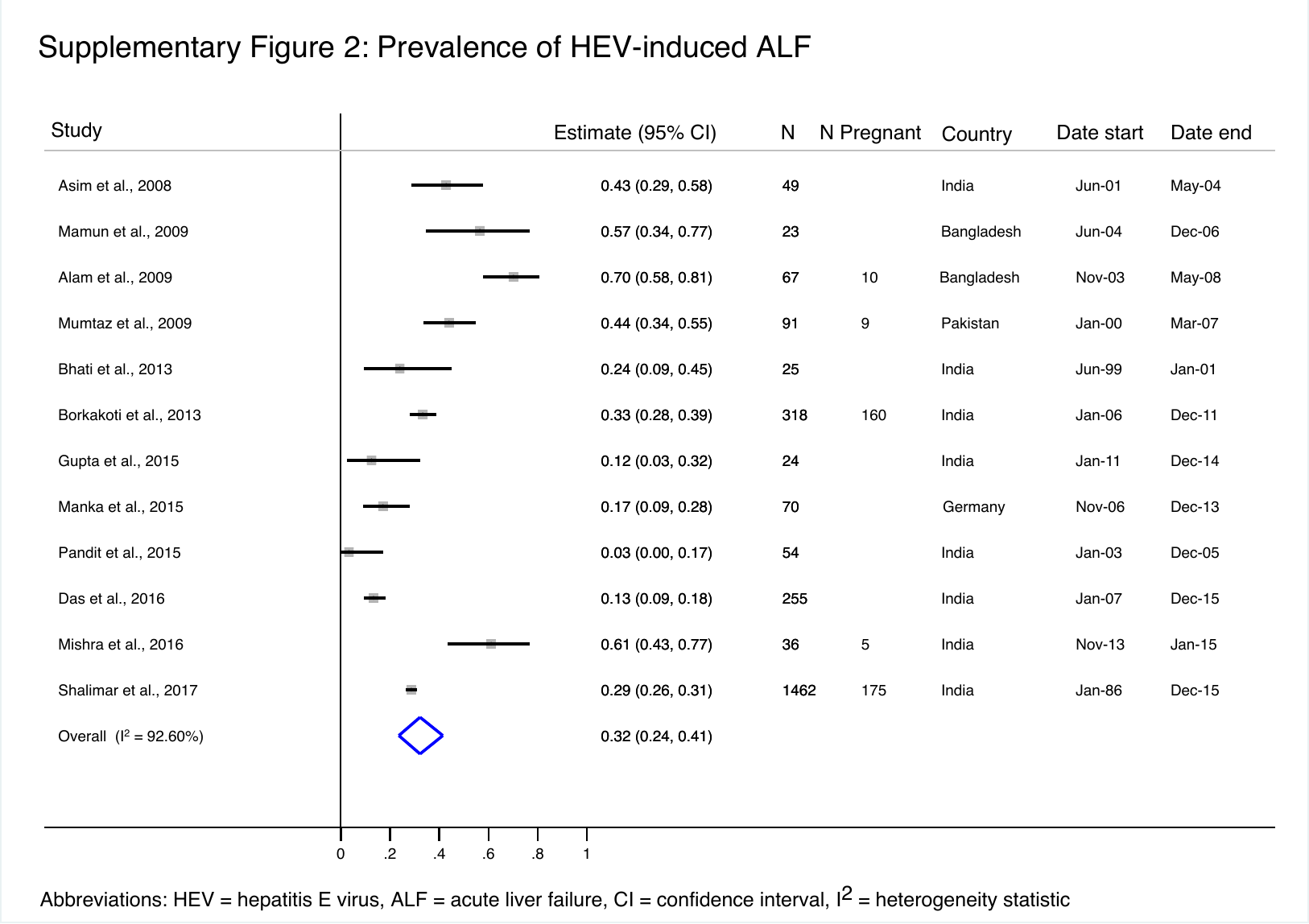

###### Supplementary Figure S3.3: Prevalence of hepatitis D virus, herpes simplex virus, cytomegalovirus, and Epstein Barr virus induced acute liver failure
